# COVID-19 mass testing: harnessing the power of wastewater epidemiology

**DOI:** 10.1101/2021.05.24.21257703

**Authors:** Stephen F. Fitzgerald, Gianluigi Rossi, Alison S. Low, Sean P. McAteer, Brian O’Keefe, David Findlay, Graeme J. Cameron, Peter Pollard, Peter T. R. Singleton, George Ponton, Andrew C. Singer, Kata Farkas, Davey Jones, David W Graham, Marcos Quintela-Baluja, Christine Tait-Burkard, David L. Gally, Rowland Kao, Alexander Corbishley

## Abstract

**Background:** COVID-19 patients shed SARS-CoV-2 RNA in their faeces. We hypothesised that detection of SARS-CoV-2 RNA in wastewater treatment plant (WWTP) influent could be a valuable tool to assist in public health decision making. We aimed to rapidly develop and validate a scalable methodology for the detection of SARS-CoV-2 RNA in wastewater that could be implemented at a national level and to determine the relationship between the wastewater signal and COVID-19 cases in the community.

**Methods:** We developed a filtration-based methodology for the concentration of SARS-CoV-2 from WWTP influent and subsequent detection and quantification by RT-qPCR. This methodology was used to monitor 28 WWTPs across Scotland, serving 50% of the population. For each WWTP catchment area, we collected data describing COVID-19 cases and deaths. We quantified spatial and temporal relationships between SARS-CoV-2 RNA in wastewater and COVID-19 cases.

**Findings:** Daily WWTP SARS-CoV-2 influent viral RNA load, calculated using daily influent flow rates, had the strongest correlation (ρ>0.9) with COVID-19 cases within a catchment. As the incidence of COVID-19 cases within a community increased, a linear relationship emerged between cases and influent viral RNA load. There were significant differences between WWTPs in their capacity to predict case numbers based on influent viral RNA load, with the limit of detection ranging from twenty-five cases for larger plants to a single case in smaller plants.

**Interpretation:** The levels of SARS-CoV-2 RNA in WWTP influent provide a cost-effective and unbiased measure of COVID-19 incidence within a community, indicating that national scale wastewater-based epidemiology can play a role in COVID-19 surveillance. In Scotland, wastewater testing has been expanded to cover 75% of the population, with sub-catchment sampling being used to focus surge testing. SARS-CoV-2 variant detection, assessment of vaccination on community transmission and surveillance for other infectious diseases represent promising future applications.

**Funding:** This study was funded by project grants from the Scottish Government via the Centre of Expertise for Waters (CD2019/06) and The Natural Environment Research Council’s COVID-19 Rapid Response grants (NE/V010441/1). The Roslin Institute receives strategic funding from the Biotechnology and Biological Sciences Research Council (BB/P013740/1, BBS/E/D/20002173). Sample collection and supplementary analysis was funded and undertaken by Scottish Water and the majority of the sample analysis was funded and undertaken by the Scottish Environment Protection Agency.

## Introduction

The COVID-19 pandemic has necessitated the rapid implementation of surveillance programmes worldwide to track and control the spread of SARS-CoV-2. Initially, such programmes relied on syndromic surveillance, community testing, contact tracing and the monitoring of morbidity and mortality rates (1-3). Community testing relies on voluntary reporting of clinical signs and is only partially able to capture the pre-symptomatic, asymptomatic and pauci-symptomatic cases of SARS-CoV-2 infection that can contribute significantly to community transmission, and are therefore subject to biases, which can influence estimates of disease burden (1, 2). Syndromic surveillance based on hospital admissions is less biased, but is subject to delays between infection and admission (2), while implementing mass swab-testing on a nationally meaningful scale is not economically feasible for most countries (2).

Early studies identified SARS-CoV-2 RNA in the faeces of infected individuals and COVID-19 has subsequently been associated with a range of gastrointestinal symptoms (4). SARS-CoV-2 has been detected in faeces from both asymptomatic and symptomatic individuals, with prolonged shedding observed up to 33 days after the initial onset of symptoms or hospitalisations (1, 4, 5). Consequentially, wastewater-based epidemiology (WBE) has been explored as a tool to track the spread of SARS-CoV-2 by many countries (1). Medema *et al*. (2020) detected SARS-CoV-2 in wastewater early in the pandemic and identified viral RNA in the wastewater of three Dutch cities and a major airport up to six days before the first reported clinical cases (6). WBE studies are now ongoing in over 50 countries (1, 7, 8). Although proving effective as a surveillance tool, understanding the impact of viral shedding dynamics in faeces, viral persistence in wastewater and wastewater flow rates on viral detection remain serious challenges. Differences between urban and rural wastewater systems and accurate normalisation to population size also must also be considered (2). Furthermore, wastewater samples are diverse and can contain PCR inhibitors; therefore, reproducible virus process controls are needed (2). Whilst there are a range of techniques for detecting viruses in wastewater, many are difficult to operationalise at a national scale. This study describes the development and implementation of a national WBE SARS-CoV-2 surveillance programme. We compared and optimised commonly used viral concentration techniques, validated Porcine Respiratory and Reproductive virus (PRRSv) as a suitable process control and optimised RT-qPCR assays for SARS-CoV-2 detection in wastewater. This methodology was adopted by the Scottish Environment Protection Agency (SEPA) and has been used to routinely monitor viral levels at 28 wastewater treatment plant (WWTP) sites across Scotland, serving 50% of the Scottish population (2.66 million people). These sites include large conurbations, as well as low-density rural and remote island communities. We demonstrate that daily SARS-CoV-2 viral RNA load can be used to predict the number of cases detected in the WWTP catchment area, with a clear statistically significant relationship observed between these two variables above site-specific case thresholds.

## Methods

### WWTP site selection

WWTP monitoring sites were selected by Scottish Water and SEPA to represent at least 50% of the population in each Scottish health board area (Table S2.1), using the minimum number of sites possible.

### Wastewater sample collection

WWTP influent was collected at each site using a refrigerated autosampler that obtained representative influent samples over each 24-hour period. Sites were typically sampled once a week, with increased frequency of sampling in response to changes in disease incidence in the community. Samples were transported and stored at 4°C prior to analysis, typically within 24-48 hours of collection.

### Wastewater concentration and detection of SARS-CoV-2

Five viral concentration methods, Methods 1 – 5, based on filtration, precipitation and adsorption were trialled (see Supplementary Materials). Method 1 was further optimised by SEPA (Method 6) and used for routine wastewater monitoring. Viral RNA was extracted from concentrated wastewater samples using commercial kits. SARS-CoV2 was detected by RT-qPCR (E-gene during method development and N1-gene during routine monitoring).

### Data collection

Two WWTP datasets were provided by SEPA via a publicly available portal (9). The first dataset reported sample date, location (WWTP name, coordinates, Health Board, and Local Authority), catchment area (CA) size (population band and population) and SARS-CoV-2 N1 and E gene average concentrations (gene copies/l). The second dataset reported the daily WWTP influent flow (l/day) and three separate N1 gene replicates for each sample. SEPA also provided the WWTP dry weather (i.e. licenced) flow (l/day) and Scottish Water the CA shapefiles for the 28 sites.

COVID-19 data in Scotland are collected by Public Health Scotland (PHS) and the dataset used in this study reports the date and location of first COVID-19 tests and first positive tests (i.e. such that ‘positivity’ is the proportion of individuals who test positive), with test results, and deaths, starting from March 1^st^, 2020. To protect patient anonymity, data were provided by PHS by “datazone”, a small-scale geographic unit identified by the National Records of Scotland (NRS) containing approximately 500 to 1000 individuals. Relevant shapefiles and population data were downloaded from the NRS portal (10), facilitating a high resolution allocation of the number of tests, detected cases (i.e. positive tests), and COVID-19 related deaths for each of the CAs.

### Data analysis

The objective was to understand the association between the concentration and daily viral RNA load of SARS-CoV-2 RNA in WWTP influent and the number of detected cases in the corresponding CA. The daily WWTP influent viral RNA load was calculated by multiplying the wastewater sample viral RNA concentration with the WWTP influent flow for the day of sampling. Since daily flow is not always available, SEPA included a flow estimate obtained with a linear regression model that considered ammonium concentration (provided by Scottish Water), catchment population, and site as independent variables (Roberts and Fang, private communication). Analyses were repeated using both reported flow rates and these estimates (see Supplementary Material).

The number of detected cases and the positive test rate were calculated by counting the number of positive and total tests over a time period ending on the day the sample was taken. This period was set to a week in the main analysis, while we tested the effect of varying it from zero days (i.e. counting only the reported cases on the day of wastewater sample collection) to 28 days on our results (see Supplementary Material).

To test the association between observed cases (*Y*_*i,j*_) and daily WWTP viral RNA load (*X*_*i,j*_), we fitted a linear mixed model:

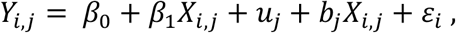

where *β*_*0*_ and *β*_*1*_ represent the fixed intercept and coefficient of the daily WWTP viral RNA load *X*_*i,j*_. Parameters *u*_*j*_ and *b*_*j*_ are the random intercept and coefficient, associated with each group *j* (catchment). Before the estimation, the dependent and independent variables were square root transformed.

We evaluated the model using the conditional pseudo-R^2^, which measures the variance explained by both fixed and random effects (11) and analysed the resulting coefficients (intercept, *β*_*0*_ + *u*_*j*_, and slope, *β*_*1*_ + *b*_*j*_) to assess the consistency of the signal and the potential causes of the differences between WWTPs. We first fitted a series of univariable linear regression models with the site’s slope or intercept as the dependent variable and population, population density, number of wastewater samples, latitude, longitude, deprivation and access indices (10) as dependent variables. We then fitted a multivariable model to each coefficient, selecting as independent variables those returning a *p*-value below 0·2 in the univariable models. Variables were then further selected through a stepwise selection in order to eliminate the statistically insignificant ones. All data manipulation and analysis was done in R 4.0.1 (12) using packages *tidyverse* (13), *lme4* (14), and *MuMIn* (15).

## Results

### Method optimisation and detection of SARS-CoV-2 RNA in WWTP influent

Reliable quantification of SARS-CoV-2 in wastewater requires consistent viral RNA extraction across a broad range of concentrations. To investigate this, aliquots of a single wastewater sample were spiked with a serial dilution of heat-inactivated SARS-CoV-2. There was no association between viral concentration and the efficiency of RNA recovery across five orders of magnitude of SARS-CoV-2 concentration (Fig S1.1.A). We next validated PRRSv (a porcine enveloped nidovirus that can be cultured *in vitro* at Containment Level 2) as a suitable surrogate process control virus for SARS-CoV-2. The extraction efficiencies of heat-inactivated SARS-CoV-2, live PRRSv and heat-inactivated PRRSv were comparable when used to spike a single wastewater sample (Fig S1.1.B). Extraction efficiencies were also comparable between SARS-CoV-2 and heat inactivated PRRSv across wastewater samples from six individual WWTPs (Fig S1.1.C). Heat-inactivated PRRSv was chosen as a process control for all subsequent testing.

Viral concentration methodologies based on filtration (Methods 1 -3), PEG precipitation (Method 4) and adsorption (Method 5) were compared. The requirement to stir larger sample volumes for 8 h made the milk powder adsorption method insufficiently scalable and so it was excluded following initial pilot trials. Filtration and PEG precipitation resulted in comparable efficiencies of PRRSv recovery, however, more variability between technical replicates was observed using PEG precipitation (Fig S1.1.D).

We compared liquid phase (influent and effluent) and solid phase (primary sludge and dewatered cake) samples for use in detection of SARS-CoV-2 RNA. Samples were taken weekly from a single plant, WWTP2, over a three-week period. Recovery of PRRSv from influent was 20% across the 3-week sample period (Fig S1.1.E), however SARS-CoV-2 RNA levels were below the limit of quantification (Fig S1.1.F).

SARS-CoV-2 RNA was detected in all primary sludge samples and 2/3 dewatered cake samples from WWTP2 despite poor recovery of PRRSv from both sludge (0·5 – 3·5%) and dewatered cake (0·2 – 0·8%). No SARS-CoV-2 RNA was detected in the effluent from WWTP2 (n=3 replicates taken weekly over 3 consecutive weeks). Although sludge and/or dewatered cake may be a more sensitive sample type for detection of SARS-CoV-2, due to sampling difficulty and differences in sludge processing methods among WWTPs, influent samples were chosen for subsequent testing. Furthermore, some WWTPs treat sludge from other sites and hence sludge may not always be representative of the WWTP CA.

Method 1 was selected to determine if SARS-CoV-2 RNA could be detected and quantified in wastewater collected from WWTPs in Scotland during the start of the pandemic. Influent samples from six wastewater treatment plants, WWTP1 – WWTP6, were tested (Fig S1.2). Samples were taken on 27^th^ March 2020, shortly before the first COVID-19 mortality peak in Scotland. A strong positive SARS-CoV-2 RNA signal of 18,000 genome equivalents per litre was detected in sample WWTP5 (Fig S1.2.A). SARS-CoV-2 RNA levels in each of the other five plants fell below our limit of quantification. Method 1 was further optimised by SEPA (Method 6; Supplementary Materials) and used for routine wastewater monitoring. Of note, detection of the N1-gene by RT-qPCR was found to be more sensitive than the E-gene (Fig S1.2.B) and therefore N1-gene detection was adopted for the national programme.

### Data analysis

The weekly number of SARS-CoV-2 reported cases, deaths and positivity are shown in Fig 1A. As of 29/1/2021, 989 wastewater samples, with three replicates each, have been analysed across 28 WWTPs, with the earliest samples taken from late May 2020 (Fig 1B). The number of samples per WWTP ranged from 12 (Stornoway, Outer Hebrides) to 112 (Shieldhall, Greater Glasgow). The CAs are distributed across Scotland (Fig 1C) and despite covering only 1·2% of Scotland’s land mass, they cover 50% of the population. Daily WWTP influent flow data was missing for 18% of the samples.

**Figure 1.**
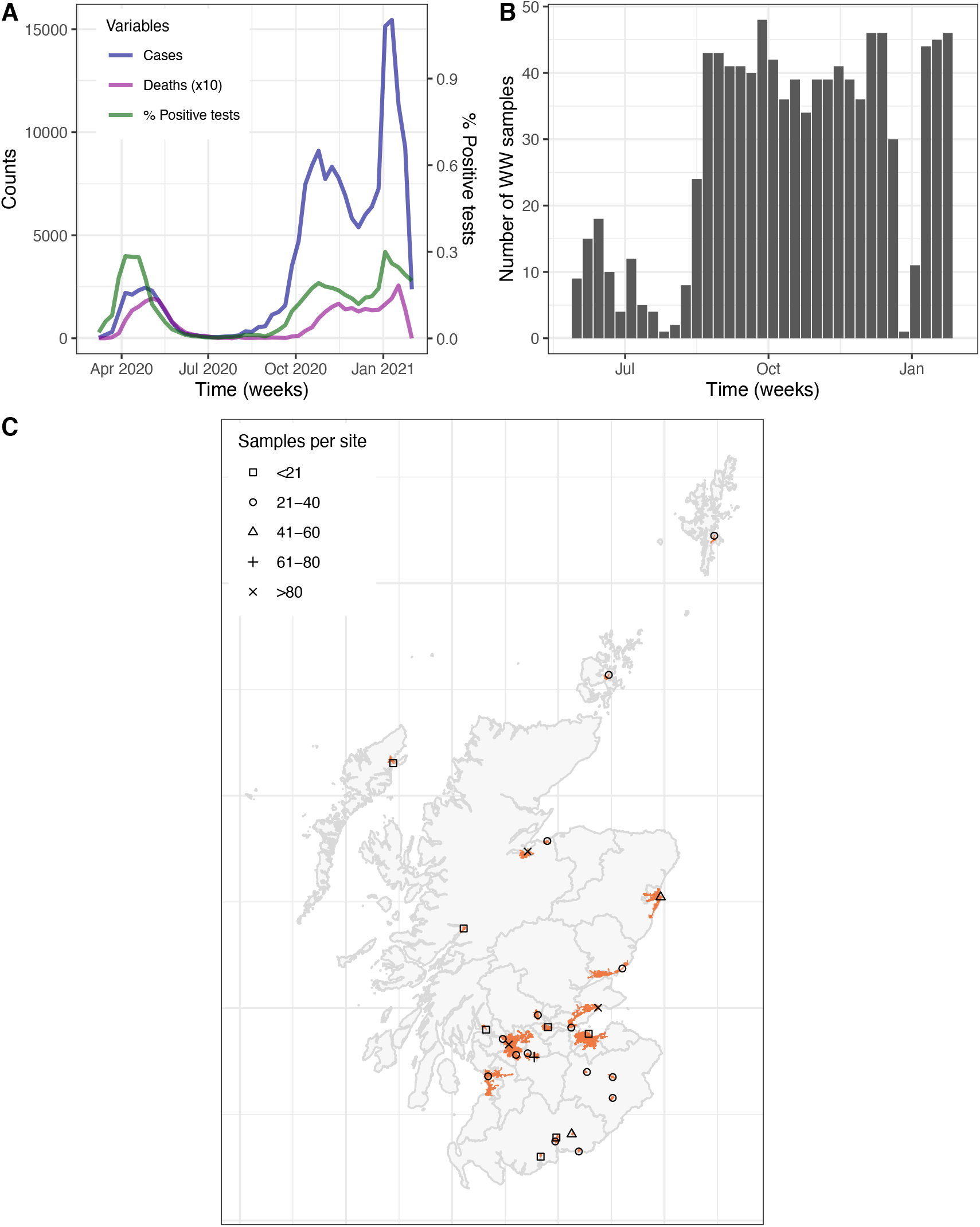
A, Number of weekly COVID-19 cases, deaths (multiplied by ten), and positive test rate in Scotland; B, weekly number of wastewater samples across the 28 study sites; C, spatial distribution of the 28 wastewater treatment plant sites with their catchment area (orange). Shape denotes the total number of samples by site (square: less than 20, circle: 21 to 40, triangle: 41 to 60, plus: 61 to 80, cross: over 80).

As evident in Fig 2, wastewater RNA viral concentration (panels A, C and E) and daily WWTP viral RNA load (panels B, D, and F) mimic the trends of the daily positive test rate (number of positive tests over the total) and the daily incidence curves, respectively. This was independent of the CA population size (Fig S2.1 to S2.5 for remaining WWTPs). Preliminary correlation analyses showed that WWTP daily viral RNA load is highly correlated with the number of COVID-19 cases detected in the CA, while the wastewater viral concentration was highly correlated with the positive test rate (Fig S2.6 and Fig S2.7). The correlations improve as the number of contributing days for case counts before sampling increases from zero to five, at which point it stabilises.

**Figure 2.**
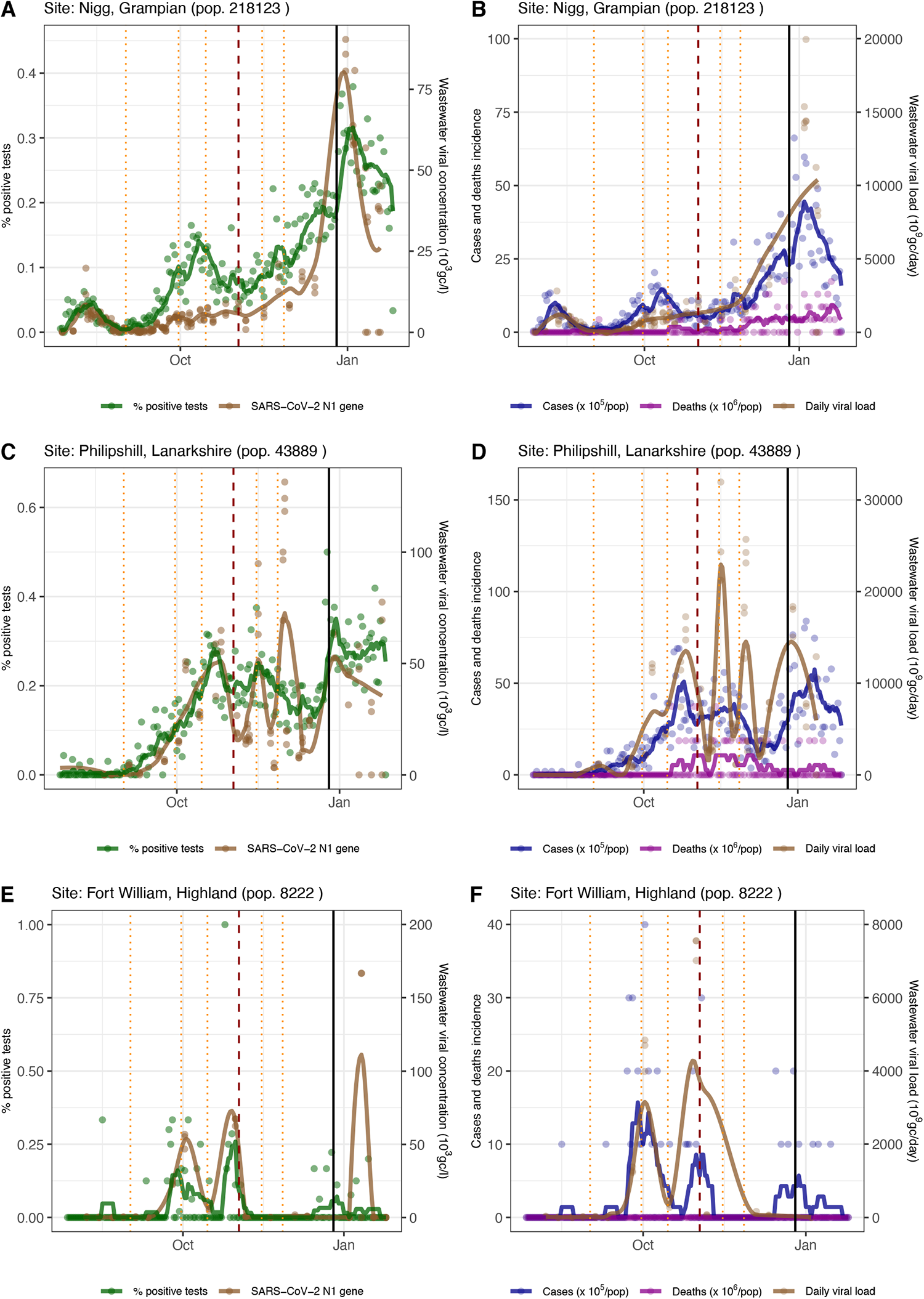
Trends of the first test positivity rate (green) and SARS-CoV-2 N1 gene concentration (brown, gc/l) in wastewater samples (panels A, C, and E); trends of COVID-19 incidence per 100,000 people (blue), deaths per 1,000,000 people (purple), and N1 gene daily load (brown, gc/day) in wastewater samples (panels B, D, and F). For positive test rate, cases, and deaths, points represent the daily value, and lines the seven-day rolling mean. For N1 gene concentration and daily load, points represent each reading of the samples, and the line was obtained by fitting a locally estimated scatterplot smoothing (LOESS) function. Data for three sites of different size are visualised here: Nigg (Grampian, panel A and B), Philipshill (Lanarkshire, panel C and D), and Fort William (Highland, panel E and F). The remaining 25 are shown in the Supplementary Material. Vertical lines mark the changes in restrictions: local or minor policy changes (orange dotted lines), the introduction of the regional tier system (dashed red line) and the post-Christmas national lockdown (black thick line).

The full mixed model explained 78% of the variance in the number of cases in the CA (conditional R^2^ = 0·78), while the daily viral RNA load as a fixed effect explained 45% of the variance (marginal R^2^ = 0·45). The null hypothesis that the sites’ random slope variance was zero, i.e. each site had the same slope gradient, was rejected with a Chi^2^ test (p ∼ 0). By varying the length of the time period used to calculate the number of cases, the conditional R^2^ ranged from 0·71 to 0·89, with an average of 0·76 across the 29 periods considered (Fig S2.8).

The mixed model fit by site is reported in Fig 3 (and Fig S2.9). While the daily WWTP viral RNA load coefficients, or slope, are an indicator of the strength of the relationship between viral RNA load and cases, the intercept provides an estimate of the limit of detected cases in each CA. The median [interquartile] estimated slope across sites was 5·2 × 10^6^ [4·50-5·37 × 10^6^], and was positive in all sites, including the confidence interval (Fig 4A). The median [interquartile] intercept was 2·01 [0·90-3·77]. The intercept varied substantially between WWTPs of different size: median 0·84 [0·63-0·90] for the smaller sites (< 10,000 population), 2·25 [1·72-3·78] for the medium-sized sites (10,000 to 100,000 population), and 5·30 [3·2-6·95] for the larger sites (> 100,000 population). This translates to a threshold of less than one recorded case from which the relationship between viral RNA load and cases is detectable in small catchments, five recorded cases in the medium-sized catchments and twenty-five cases in the large catchments. Among the latter group, Dalmuir and Meadowhead were outliers, with higher intercept and lower slope compared with similar-sized catchments (Fig 4C).

**Figure 3.**
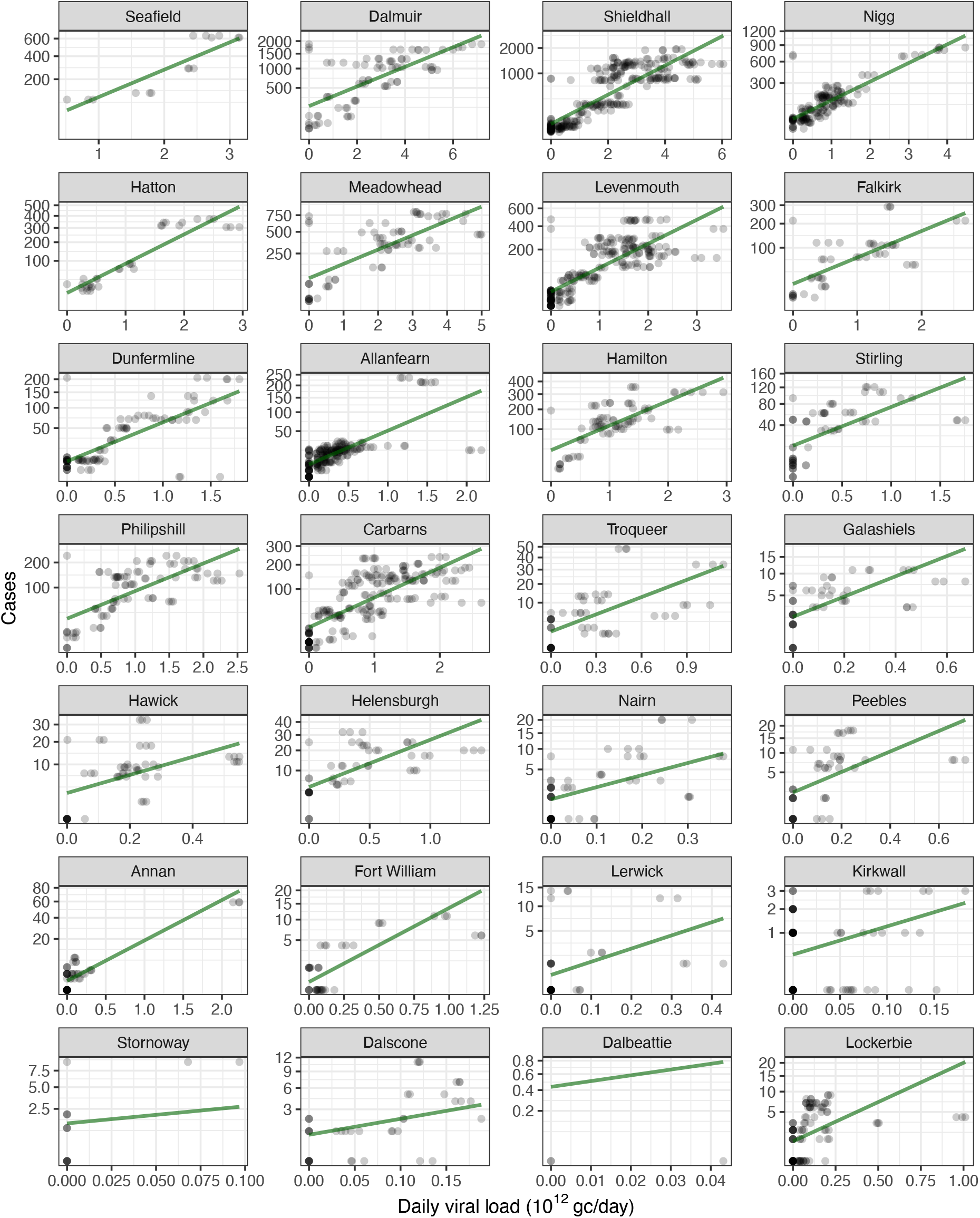
Linear regression mixed model fit for the 28 wastewater treatment plants, ordered by their catchment population size. Each WWTP regression is plotted with independent axes limits, see Figure S.2.8. for a version of the plot with fixed axes.

**Figure 4.**
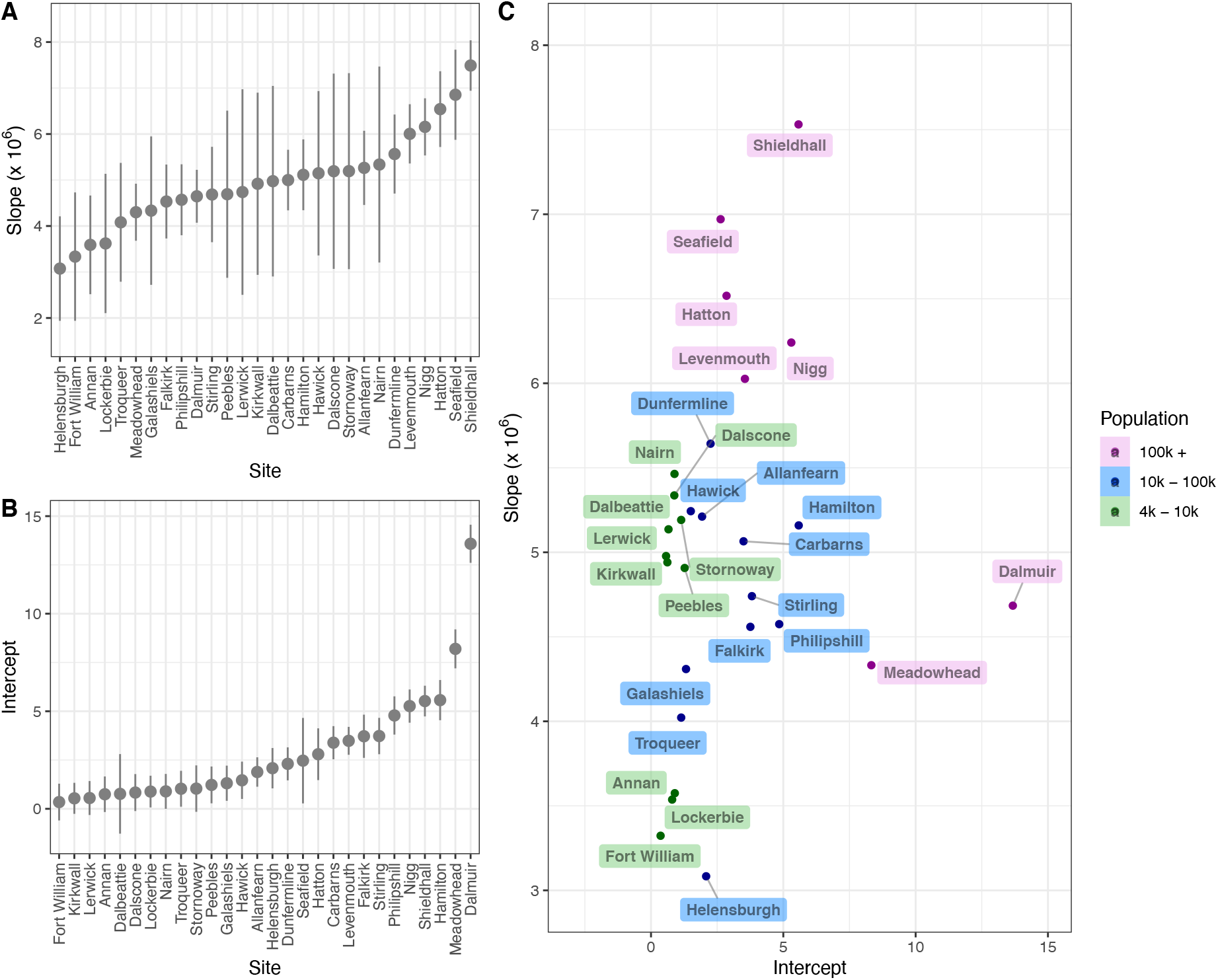
Linear mixed model coefficients: slopes (panel A) and intercept (panel B), ordered by coefficient size. Points correspond to the mean and bars correspond to confidence interval. Panel C shows the relationship between slope and intercept, with points and labels coloured by catchment population size.

The variables that best explain differences in mixed model slopes across WWTPs were the population size and the number of samples taken, although geographical longitude (not significant) was retained after multivariable model stepwise selection (Table 1). The CA population size and deprivation index were significant in explaining the differences in the mixed model intercepts (see Fig S2.10 for single variable plots).

**Table 1.** Results of the univariable and multivariable linear models to determine the variables that influence the mixed model slope and intercept for different sites. Deprivation and access indices measure the relative deprivation and the access to healthcare services respectively of a datazone. They were included as potential causes of bias in case detection. The R^2^ of the two multivariable linear models was 0.45 for the slope, and 0.50 for the intercept (both *p* < 0.001).

| Dependent variable | Independent variable | Univariable linear model |  | Multivariable linear model |  |
| --- | --- | --- | --- | --- | --- |
|  |  | Coefficient | *p* | Coefficient | *p* |
| Mixed model groups slopes | *Number of samples* | **0·42** | **0·012** | **0·30** | **0·042** |
|  | ***Population*** | **0·56** | **<0·001** | **0·47** | **0·003** |
|  | *Density* | 0·27 | 0·129 | (dropped by stepwise selection) |  |
|  | Latitude | 0·15 | 0·435 | - | - |
|  | *Longitude* | 0·31 | 0·179 | 0·28 | 0·109 |
|  | Deprivation index | 0·15 | 0·373 | - | - |
|  | *Access index* | -0·44 | 0·007 | (dropped by stepwise selection) |  |
|  | *Multivar. intercept* | - | - | 0·15 | 0·132 |
| Mixed model groups intercepts | Number of Samples | 0·20 | 0·236 | - | - |
|  | ***Population*** | **0·53** | **<0·001** | **0·45** | **0·002** |
|  | *Density* | 0·48 | 0·001 | (dropped by stepwise selection) |  |
|  | Latitude | -0·16 | 0·370 | - | - |
|  | Longitude | -0·25 | 0·242 | - | - |
|  | ***Deprivation index*** | **0·37** | **0·009** | **0·27** | **0·030** |
|  | *Access index* | -0·42 | 0·005 | (dropped by stepwise selection) |  |
|  | *Multivar. intercept* | - | - | 0·01 | 0·908 |

## Discussion

SARS-CoV-2 WBE has rapidly become an important surveillance tool for COVID-19 around the world. This work uniquely describes the establishment of a WBE programme covering 50% of a country’s population across a wide range of WWTP sizes, including large cities and small remote rural and island communities. We have used granular geospatial data to determine accurate estimates of recorded COVID-19 cases within each CA. We demonstrate the existence of a strong and measurable statistically significant relationship between the SARS-CoV-2 daily WWTP viral RNA load and the number of detected cases in the week preceding wastewater sample collection.

The precision of this estimate varies between sites, with differences in the slope mostly attributed to the size of the population being served. Our results identified a stronger relationship between cases and viral RNA load in the larger WWTPs. The identified threshold for detection was typically under 25 cases, and for some smaller WWTPs, a single detected community case was sufficient to yield a positive wastewater result. Compared to similar-sized WWTPs, Meadowhead and Dalmuir were outliers (Fig 4C); given their size, the slopes imply a poorer relationship between detected cases and WWTP daily viral RNA load, and intercepts a poorer sensitivity than expected. These WWTPs are defined by fragmented and highly dispersed CAs compared to most WWTPs of this size. Thus network architecture may be important, and sub-catchment sampling may be necessary for large, fragmented, and/or dispersed networks. Deprivation also had a significant impact on the intercept, possibly due to differences in case reporting and/or viral RNA load per case. Combined, these factors meant that the limit of detection of cases per 100,000 population was highly variable between WWTPs: median 6·5 [4·6-10·7] for the smaller sites, 19·0 [8·0-27·9] for the medium-sized sites, and 10·3 [6·0-23·0] for the larger sites.

Studies in other countries have identified similar trends in SARS-CoV-2 levels in wastewater and COVID-19 cases in the CA, including in the USA (16), Australia (17), France (18), and Spain (19). Importantly, we demonstrate how WBE can be adopted across a range of catchments, from densely populated urban areas (Edinburgh and Glasgow), to smaller towns, rural areas and islands. In contrast to previous studies, we demonstrate the value of obtaining flow measurements from WWTPs to calculate daily viral RNA loads, which display a stronger correlation with detected community case numbers, compared with viral concentration data alone (Fig S2.6). This strong correlation demonstrates that degradation of viral RNA within the wastewater network is minimal over the considered time scale, as reported by other studies (20).

Our typically low limits of detection show that wastewater surveillance can be particularly valuable for areas reaching low prevalence and is therefore suitable as a logistically sustainable and cost-effective early warning system, making a targeted community testing strategy viable. For WWTPs collecting wastewater from cities, it is harder to isolate small clusters of infections. This hurdle can be overcome by sampling a site “upstream” to the WWTP (i.e. within the sewerage network) to improve spatial resolution. This is currently taking place in Scotland, with local health boards using sub-catchment wastewater sampling to direct surge testing.

For smaller catchments, the size and the spatial resolution is already fine enough to inform community interventions, however a potential issue here is the variability in the signal. Specifically, we observed sudden spikes in the viral RNA load or viral concentration in many small WWTPs (Fig 2, E and F; Fig S2.4; Fig S2.5). While smaller catchments might be more sensitive to individual variations in shedding, these spikes might also be caused by one or two households being infected in a short period of time. Given the sensitivity of these smaller WWTPs to a small number of cases, this may explain these sudden variations in the SARS-CoV-2 daily viral RNA load.

Whilst we have shown that daily viral RNA load has the best correlation with detected cases, daily WWTP flow measurements are not always available. This may be more of a problem in smaller WWTPs, where flow rates regularly exceed the working range of the flow meter or in low resource settings, however our model retained substantial detection power when daily flow was estimated using easily obtained ammonium concentrations, with the conditional R^2^ dropping by only 2% (R^2^= 0·76).

To better understand the relationship between WWTP viral RNA load and infected individuals, we need to consider the level of viral shedding in faeces and how this varies over time. Whilst SARS-CoV-2 RNA can be detected in the faeces of hospitalised patients for over four weeks (21, 22), our work and that of others (23) implies a relatively short period of time over which infected individuals substantially contribute to the wastewater signal. This was observed in two distinct sensitivity analyses, one on correlations and the other on mixed model performance (see Supplementary material). Specifically, the correlation between cases and viral RNA load (and between positive test rate and viral concentration) stabilises once detected cases are included up to and including the five days prior to wastewater sampling. Furthermore, even with declining incidence, when the cumulative effect of older infections would be expected to have a greater contribution to the overall signal if shedding duration was long, the conditional R^2^ of the mixed models did not deteriorate significantly (0·76 compared to 0·78 when incidence was increasing), and was consistent with a short period of peak viral shedding. Unfortunately, there is currently very limited data on faecal shedding of SARS-CoV-2 RNA in non-hospitalised individuals. Our understanding of the relationship between the WWTP viral RNA load and infected individuals is further complicated by the biases in community testing and movement (although restricted during lockdowns) of individuals between CAs. Specifically, testing of symptomatic individuals is unlikely to fully reflect the population incidence, with an analysis of English data suggesting that approximately 1 in 4 cases were being reported via community testing up to November 2020 (24).

The value of our results extends beyond the first year of the COVID-19 pandemic. We have demonstrated how COVID-19 WBE can be implemented at a national scale across a diverse range of urban and remote communities. At the time of writing, this programme has been expanded to cover 75% of the population of Scotland and is being used by local health boards to direct surge testing within the community. This programme will continue to be important during the rollout of COVID-19 vaccinations, particularly with respect to disclosing areas of on-going disease transmission and surveillance for novel SARS-CoV-2 variants (25, 26). It also provides public health authorities with an unbiased surveillance network for other viral and bacterial infections, including antimicrobial resistance genes, shed in faeces. Until the COVID-19 pandemic, WBE was predominantly limited to the surveillance of a narrow range of viruses (e.g. polio, norovirus, Hepatitis A/E) in low resource, sewered settings (27-29). This study demonstrates the rapid inception, development, validation and operationalisation of a national COVID-19 WBE programme to provide highly cost-effective community surveillance during the pandemic.

## Data Availability

All scripts and models used in this study will be made available upon request to the corresponding authors. Wastewater SARS–CoV–2 RNA levels are publicly available from SEPA here: https://informatics.sepa.org.uk/RNAmonitoring/. The authors cannot share the wastewater treatment plant catchment areas and patient data aggregated at the datazone level, which are subject to data sharing agreements and must be requested directly from Scottish Water and Public Health Scotland respectively.

## Acknowledgements

The authors would like to thank all the sampling and courier staff at Scottish Water and the team of scientists and other staff at the Scottish Environment Protection Agency for their efforts in acquiring, transporting and analysing the wastewater samples throughout the pandemic, as well as the electronic Data Research and Innovation Service (eDRIS) who provided the COVID-19 test and death data. We would also like to acknowledge the support of Andrew Millar, the Scottish Government’s Chief Scientific Adviser for Environment, Natural Resources and Agriculture.

