## Supplementary material for "COVID-19 mass testing: harnessing the power of wastewater epidemiology"

### Supplementary Methods

#### Wastewater concentration

Method 1. A 20 ml or 40 ml aliquot was spiked with 15 µl of spike virus. Samples were clarified by centrifugation to pellet large debris (4000 x *g*, 30 min, 4°C) followed by filtration using a 0.45 µm syringe filter (Fisher). Samples were concentrated to 500 µl by centrifugation using Vivaspin 20 centrifugal filters with a 50 kDa MWCO (Sartorius) and further concentrated to 140 – 200 µl using Amicon® Ultra 0.5 ml centrifugal filters with a 3 kDa MWCO (Millipore).

Method 2. A 20 ml aliquot was spiked with 15 µl of spike virus. Samples were clarified by centrifugation to pellet large debris (4000 x *g*, 30 min, 4°C) followed filtration using a 0.45 µm syringe filter (Fisher). Samples were concentrated to 140 µl by centrifugation using Vivaspin 20 centrifugal filters with a 50 kDa MWCO (Sartorius).

Method 3. A 20 ml aliquot was spiked with 15 µl of spike virus. Samples were clarified by centrifugation (10,000 x *g*, 30 min, 4°C). Samples were concentrated to 500 µl by centrifugation using Vivaspin 20 centrifugal filters with a 50 kDa MWCO (Sartorius) and further concentrated to 140 – 200 µl using Amicon® Ultra 0.5 ml centrifugal filters with a 3 kDa MWCO.

Method 4. A 20 ml aliquot was spiked with 15 µl of spike virus. Samples were clarified by centrifugation to pellet large debris (4000 x *g*, 30 min, 4°C). Sample pH was adjusted to 7 – 7.5 if necessary. Polyethylene glycol 8000 (PEG8000) (8% w/v) and NaCl (1.8% w/v) were added to each sample and mixed until PEG was no longer visible. Viral precipitation was allowed to proceed overnight at $4^0$C before samples were centrifuged at 10,000 x *g* for 30 min at $4^0$C. Supernatant was discarded and PEG pellets containing precipitated virus were dissolved in distilled water.

Method 5. One litre wastewater samples were adjusted to pH 3.5 with 1 M HCl and acidified 1 % w/v skimmed milk powder solution acidified to pH 3.5 with HCl was added (0.01% w/v final concentration). Samples were incubated with stirring at room temperature for 8 h to allow for adsorption of viral particles. 950 ml of supernatant was removed by pipetting the remaining precipitated skimmed milk powder solution containing adsorbed viral particles was harvested by centrifugation (4,500 x *g*, 1 h, 12 °C). Supernatant was removed and concentrated pellets were dissolved in 3 ml 0.2 M phosphate buffer (pH 7.5). Samples were treated with 6 ml 0.25 M glycine buffer (pH 9.5) for 45 min before an additional 10 ml phosphate buffer was added, and ultracentrifugation was performed at 4,500 × *g* for 20 min at 12 °C. Resulting pellets were resuspended in 200 µl phosphate buffer (pH 7.5).

Method 6 (SEPA): For each wastewater sample, 2 x 50 ml duplicates were spiked with a known quantity of PRRSv sample process control. Samples were clarified by centrifugation to pellet large debris (4000 x g, 10 min, 4°C) and supernatants were transferred to a clean tube taking care not to disturb the pellet. 15 ml of supernatant was concentrated by centrifugation (4000 x g, 10 min, 4°C) using Amicon/Centricon centrifugal filters with a 50 kDa MWCO (MerckMillipore). Flow-through was discarded, filters were refilled with supernatant and samples were again concentrated by centrifugation. Concentration steps were repeated until each 50 ml of supernatant was concentrated to <250 uL. Concentrated sample was transferred to nuclease free 2 ml Eppendorf tube and used for RNA extraction.

**RNA extraction**

Viral RNA was extracted from 140 – 250 µl concentrated wastewater samples using the QiAmp viral RNA extraction kit (Qiagen) according to the manufacturer's guidelines and eluted in a final volume of 60 µl elution buffer. To avoid PEG contamination, RNA was extracted from samples processed by Method 4 using Trizol reagent (Fisher). RNA pellets were dissolved in 60 µl QiAmp elution buffer. For solid phase sludge and dewatered cake samples, RNA was extracted directly from 2 g of sample using the RNeasy PowerSoil Total RNA kit (Qiagen) according to the manufacturer's guidelines.

**RT-qPCR**

E-gene amplification was used for detection of SARS-CoV-2 for all method development assays and N1-gene amplification was used by SEPA for nationwide monitoring. Primers and probes targeting the N- and E-genes of the SARS-CoV-2 genome used in this study are listed in Table S1.1. For method development, one-step multiplex RT-qPCR reactions were carried out using Reliance One-Step Multiplex RT-qPCR supermix (Bio-Rad) according to the manufacturer's guidelines. Each 20 µl reaction contained 6 µl RNA template and primer and probes were used at a final concentration of 500 nM and 200 nM, respectively. RT-qPCR was carried out using a CFX96 Touch real-time PCR detection system (Bio-Rad) with the following cycling conditions: 50°C for 10 min (1-cycle), 95°C for 10 min (1-cycle), 95°C for 10 s followed by 60°C for 30 s (40-cycles).

One-step RT-qPCR reactions were carried by SEPA using Luna® Universal One-Step RT-qPCR Kit (NEB) according to the manufacturer's guidelines. Each 20 µl reaction contained 6 µl RNA template and primer and probes were used at a final concentration of 500 nM and 200 nM, respectively. RT-qPCR was carried out using a AriaMx Real-time PCR System (Agilent) with the following cycling conditions: 55°C for 10 min (1-cycle), 95°C for 1 min (1-cycle), 95°C for 10 s followed by 55°C for 30 s (45-cycles).

The nCoV-ALL-control plasmid (Eurofins) was used to generate standard curves for quantification of SARS-CoV-2. For quantification of PRRSv, a fragment of the Orf7 gene was amplified using the control primer set (Table S1.1) and then cloned into plasmid pJET2.1 using a CloneJet™ PCR cloning kit (Thermofisher) according to the manufacturer's guidelines. Plasmid pJET2.1 containing PRRSv Orf7 was used to generate standard curves for quantification of PRRSv. RT-qPCR results were only considered if no amplification occurred in no template controls (NTC) and standard curves met the following criteria: an $R^2$ value > 0.9, slope (y) was between -3.1 and -3.92 and efficiency was between 80 and 110%. The limit of quantification was defined as the most dilute qPCR standard i.e. 6 copies per µl, equivalent to 1,500 genome equivalents per litre when 40 ml wastewater was processed. The NTC was negative (i.e. no Ct determined) for all qPCR assays.

**Preparation of viral spikes**

Porcine respiratory and reproductive syndrome virus (PRRSv) was cultured and harvested as described previously (9). Briefly, field isolates of the virus were isolated and passaged on primary porcine alveolar macrophages (PAMs). Viral load was quantified by RT-qPCR against DNA and RNA standards as well as TCID50 endpoint titration. SARS-CoV-2 was isolated from oronasal patient swabs obtained through the NHS Lothian Biobank. Patient samples were diluted 1:1 in DMEM containing 100IU/ml penicillin and 10 g/ml streptomycin (Invitrogen; P/S), filtered through a 0.1 µm syringe filter, and a confluent layer of Vero E6 cells inoculated. After 1.5 h, medium was replaced by DMEM with P/S and 10% fetal calf serum (HI, GE Healthcare). Virus sequences were compared against the patient isolate after each passage using the ARTIC Oxford Nanopore MinION sequencing protocol (v2 at the time of use for this project). Viral load was quantified by RT-qPCR against DNA and RNA standards as well as TCID50 endpoint titration. Culture supernatants were used as a process control viral spike. PRRSv and SARS-CoV-2 culture supernatants were heat-inactivated in a validated protocol at 70°C for 10 min using thin-wall 0.2 ml tubes in a PCR machine to ensure core temperature.

**Table S1.1.** RT-qPCR primer and probe sequences.

| | Forward primer | Reverse primer | Probe |
|---|---|---|---|
| E-Sarbeco | ACAGGTACGTTAATAGTTAATAGCGT | ATATTGCAGCAGTACGCACACA | YAKYE-ACACTAGCCATCCTTACTGCGCTTCG -BHQ1 |
| 2019-nCoV-N1 | GACCCCAAAATCAGCGAAAT | TCTGGTTACTGCCAGTTGAATCTG | FAM-ACCCCGCATTACGTTTGGTGGACC -BHQ1 |
| Control (PRRSV1) | CAGGACTTCGGAGCCTCGT | AGCAACTGGCACAGTTGATTGA | Cy5-ACGAGCTGTTAAACGAGGA -3IAbRQSp |

YAKYE = Yakima Yellow RED = Texas Red BHQ = Black hole quencher 3IAbRQSp=Iowa Black

Primers were selected on the basis of findings circulated in a preprint by Vogels et al in April 2020 (doi: https://doi.org/10.1101/2020.03.30.20048108), subsequently published in July 2020: Vogels, C.B.F., Brito, A.F., Wyllie, A.L. et al. Analytical sensitivity and efficiency comparisons of SARS-CoV-2 RT–qPCR primer–probe sets. Nat Microbiol 5, 1299–1305 (2020). https://doi.org/10.1038/s41564-020-0761-6

### Supplementary Results – Section 1

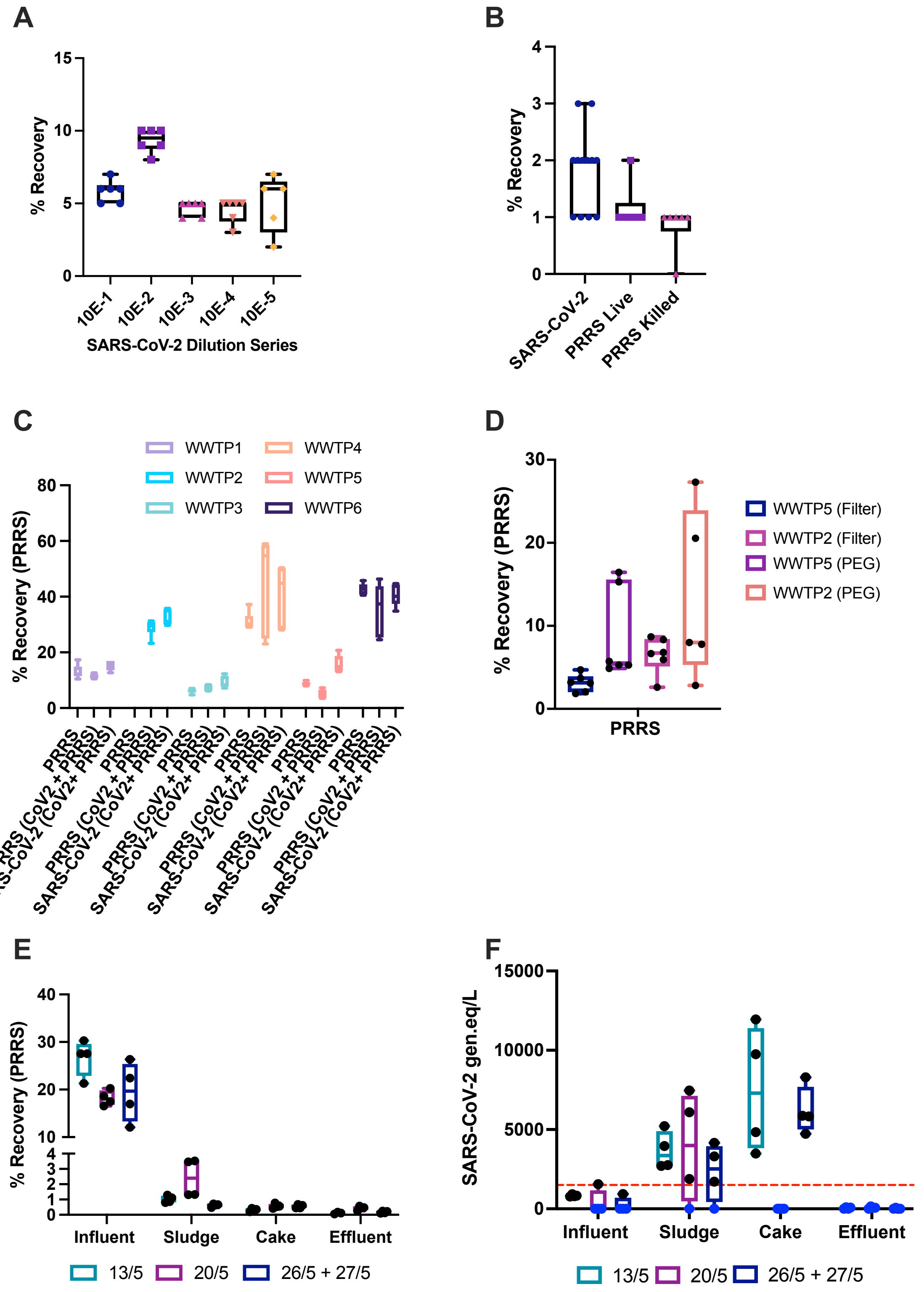

**Fig S1.1.** Method optimisation for SARS-CoV-2 concentration and detection from wastewater (a) Recovery of serially diluted heat-inactivated SARS-CoV-2 from sample WWTP3. Viral recovery was quantified for 10-fold serially diluted heat-inactivated SARS-CoV-2 over five orders of magnitude. (b) Comparison of viral recoveries for heat-inactivated SARS-CoV-2 (blue), live PRRSv (magenta) and heat-inactivated PRRSv (pink) when used to spike a single wastewater sample. (c) Comparison of spike viral recoveries from six different WWTPs, WWTP1 – WWTP 6. Samples were spiked with either heat-inactivated PRRSv alone or double spiked with heat-inactivated PRRSv and heat-inactivated SARS-CoV-2. Recoveries for PRRSv from both single and double spiked WW samples and SARS-CoV-2 are shown. Note higher recovery of SARS-CoV-2 from WWTP5 compared to PRRSv due to pre-existing SARS-CoV-2 in this sample prior to spiking. (d) Comparison of PRRSv spike recovery from WWTP2 (Pink and Orange) and WWTP5 (Blue and Purple) using either filtration (Method 1) or PEG precipitation (Method 4) for virus concentration. (e) PRRSv spike recovery from influent, primary sludge, dewatered cake and effluent samples from WWTP3. Samples were taken over a three-week period in May 2020: 13$^{th}$ May (Teal), 20$^{th}$ May (Purple) and 26$^{th}$ or 27$^{th}$ May (Navy). (f) Absolute quantification of SARS-CoV-2 present in influent, sludge, dewatered cake and effluent samples from WWTP3. Samples were taken over a three-week period in May 2020: 13$^{th}$ May (Teal), 20$^{th}$ May (Purple) and 26$^{th}$ or 27$^{th}$ May (Navy). SARS-CoV-2 genome equivalents per litre wastewater are shown. Red dotted line indicates the limit of quantification by RT-qPCR. Data points below this line were either indistinguishable from the NTC (blue) or had a Ct value greater than the most dilute standard (black). Data is derived from a minimum three (a – d) or two (e – f) biological replicates with two technical replicates of each used for RT-qPCR. Box and whisker plots showing min and max values are shown for all data. Gen.eq/L = genome equivalents per litre.

A

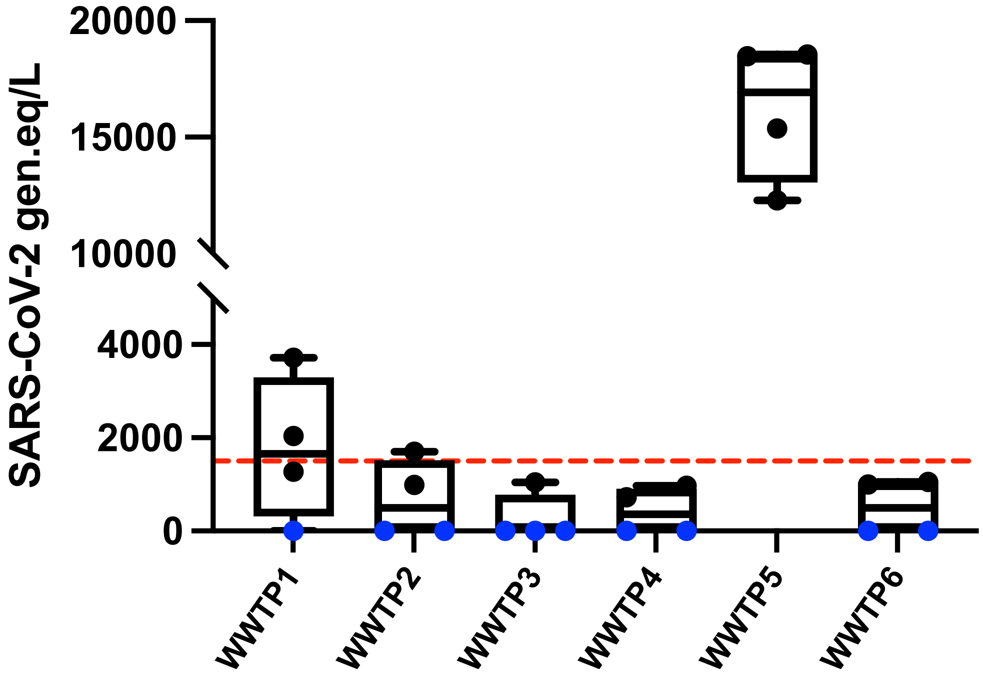
SARS-CoV-2 gen.eq/L
20000
15000
10000
4000
2000
0
WWTP1
WWTP2
WWTP3
WWTP4
WWTP5
WWTP6

B

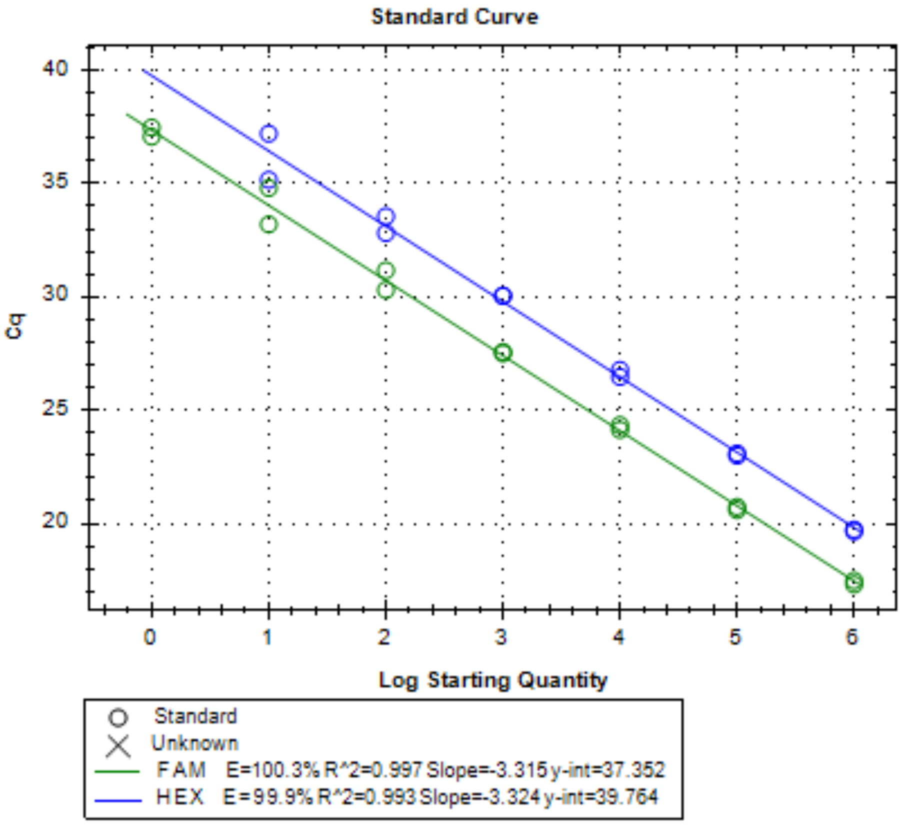
Standard Curve
Cq
40
35
30
25
20
0
1
2
3
4
5
6
Log Starting Quantity
Standard
Unknown
FAM E=100.3% R^2=0.997 Slope=-3.315 y-int=37.352
HEX E=99.9% R^2=0.993 Slope=-3.324 y-int=39.764

**Fig S1.2.** (a) Absolute quantification of SARS-CoV-2 present in wastewater in Scotland during the COVID-19 pandemic. SARS-CoV-2 RNA present in samples from six WWTPs (WWTP 1 – WWTP 6) was quantified by RT-qPCR. The treatment plants sampled were geographically distinct and served significant proportions of known Scottish NHS health board regions. SARS-CoV-2 genome equivalents per litre wastewater (gen.eq/L) are shown. Red dotted line indicates the limit of quantification by RT-qPCR. Data points below this line were either indistinguishable from the NTC (blue) or had a Ct value greater than the most dilute standard (black). Data is derived from two biological replicates with two technical replicates of each used for RT-qPCR. Box and whisker plots showing min and max values are shown for all data. (b) Comparison of N and E genes for use in RT-qPCR. SARS-CoV-2 primer/probe pairs targeting the N1 gene (FAM) and E gene (HEX) were compared by qPCR using the nCoV-ALL plasmid across a seven point tenfold serial dilution as a standard curve template. Quantification cycles (Cq) and corresponding log starting quantities of plasmid template are shown for two technical replicates at each dilution.

### Supplementary Results – Section 2

**Table S2.1.** List of Scottish wastewater treatment plants (WWTPs) included in the study and information for the catchment areas (CAs).

| Wastewater treatment plant | Health board | Local authority | Site dry weather flow ($m^3$/day) | Catchment area ($km^2$) | Population | Population density (people/$km^2$) | SARS-CoV-2 samples |
|---|---|---|---|---|---|---|---|
| Seafield | Lothian | City of Edinburgh | 261450 | 193.9 | 605,569 | 3,123 | 20 |
| Dalmuir | Greater Glasgow and Clyde | West Dunbartonshi re | 217500 | 123.2 | 428,173 | 3,475 | 30 |
| Shieldhall | Greater Glasgow and Clyde | Glasgow City | 273024 | 109.3 | 376,856 | 3,448 | 112 |
| Nigg | Grampian | Aberdeen City | 77006 | 72 | 218,123 | 3,029 | 60 |
| Hatton | Tayside | Angus | 71400 | 68.4 | 194,131 | 2,838 | 28 |
| Meadowhead | Ayrshire and Arran | North Ayrshire | 85782 | 84.1 | 190,898 | 2,270 | 32 |
| Levenmouth | Fife | Fife | 88500 | 49.6 | 116,205 | 2,343 | 87 |
| Falkirk | Forth Valley | Falkirk | 28400 | 28.5 | 70,966 | 2,490 | 18 |
| Dunfermline | Fife | Fife | 25000 | 25.6 | 69,986 | 2,734 | 34 |
| Allanfearn | Highland | Highland | 25000 | 32.1 | 62,058 | 1,933 | 89 |
| Hamilton | Lanarkshire | South Lanarkshire | 20223 | 15.1 | 51,156 | 3,388 | 28 |
| Stirling | Forth Valley | Stirling | 26100 | 16.5 | 48,428 | 2,935 | 22 |
| Philipshill | Lanarkshire | South Lanarkshire | 16898 | 16.7 | 43,889 | 2,628 | 31 |
| Carbarns | Lanarkshire | North Lanarkshire | 17129 | 16.6 | 42,545 | 2,563 | 73 |
| Troqueer | Dumfries and Galloway | Dumfries and Galloway | 13865 | 13.6 | 24,294 | 1,786 | 24 |
| Galashiels | Borders | Scottish Borders | 8613 | 5.3 | 14,921 | 2,815 | 22 |
| Hawick | Borders | Scottish Borders | 4500 | 5.2 | 14,679 | 2,823 | 24 |
| Helensburgh | Highland | Argyll and Bute | 3788 | 5.3 | 12,814 | 2,418 | 20 |
| Nairn | Highland | Highland | 3560 | 4.6 | 9,729 | 2,115 | 21 |
| Peebles | Borders | Scottish Borders | 3693 | 3.2 | 8,352 | 2,610 | 24 |
| Annan | Dumfries and Galloway | Dumfries and Galloway | 4409 | 2.9 | 8,256 | 2,847 | 21 |
| Fort William | Highland | Highland | 4598 | 3.6 | 8,222 | 2,284 | 18 |
| Lerwick | Shetland | Shetland Islands | 3855 | 3.5 | 7,843 | 2,241 | 25 |
| Kirkwall | Orkney | Orkney Islands | 3303 | 4.5 | 7,757 | 1,724 | 40 |
| Stornoway | Western Isles | Na h-Eileanan an Iar | 4582 | 5.8 | 7,247 | 1,249 | 12 |
| Dalscone | Dumfries and Galloway | Dumfries and Galloway | 2326 | 3.5 | 5,548 | 1,585 | 16 |
| Dalbeattie | Dumfries and Galloway | Dumfries and Galloway | 3240 | 2.4 | 4,227 | 1,761 | 16 |
| Lockerbie | Dumfries and Galloway | Dumfries and Galloway | 2144 | 1.8 | 4,128 | 2,293 | 42 |
| Total sampled catchments | | | - | 917 | 2,657,000 | 2,898 | 989 |
| Total Scotland | | | - | 77,933 | 5,313,600 | 697 | - |
| Percentage (%) | | | - | 1.2 | 50.0 | - | - |

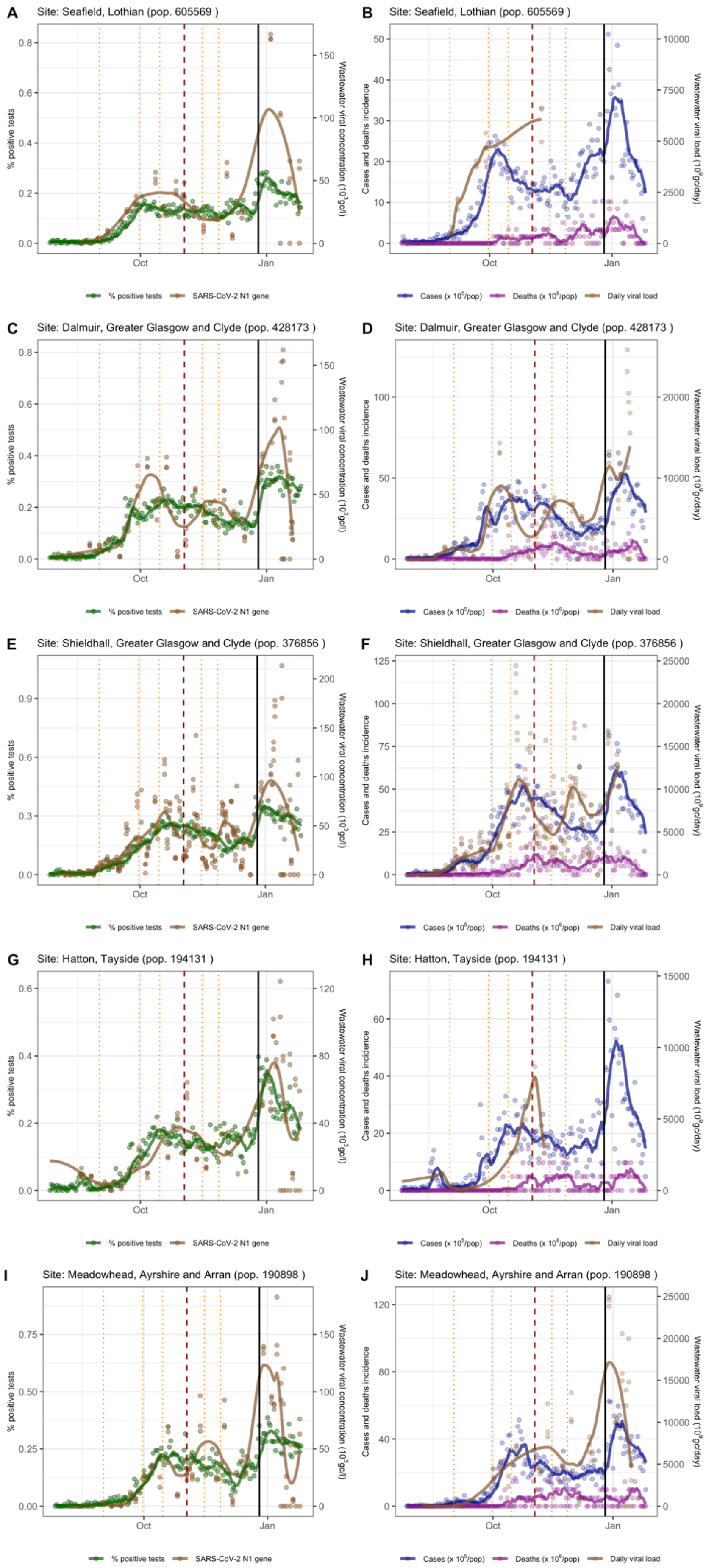


**Figure S2.1**. Trends of the first test positivity rate (green) and SARS-CoV-2 N1 gene concentration (brown, gc/l) in wastewater samples (panels A, C, E, G, and I); trends of incidence per 100,000 people (blue), deaths per 1,000,000 people (purple), and N1 gene daily viral load (brown, gc/day) in wastewater samples (panels B, D, F, H, and J). For first test positivity rate, cases, and deaths, dots represent the daily value, and lines the seven-day rolling mean. For N1 gene concentration and daily viral load, dots represent each reading of the samples, and the line was obtained by fitting a LOESS. Five wastewater treatment sites are reported: Seafield (A and B), Dalmuir (C and D), Shieldhall (E and F), Hatton (G and H), and Meadowhead (I and J). Vertical lines mark the changes in restrictions: local or minor policy changes (orange dotted lines), the introduction of the regional tier system (dashed red line) and the post-Christmas (26/12/20) national lockdown (black thick line).

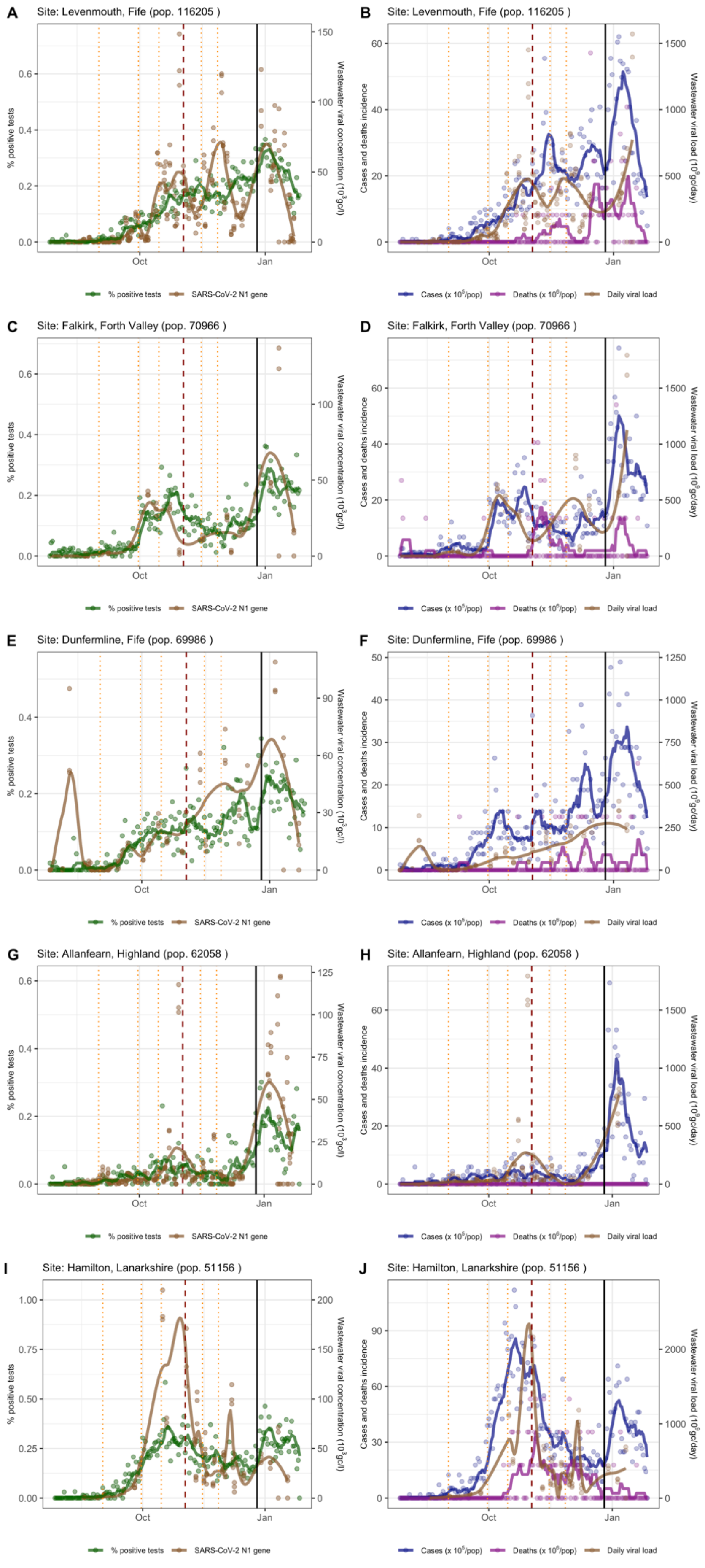


**Figure S2.2**. Trends of the first test positivity rate (green) and SARS-CoV-2 N1 gene concentration (brown, gc/l) in wastewater samples (panels A, C, E, G, and I); trends of incidence per 100,000 people (blue), deaths per 1,000,000 people (purple), and N1 gene daily viral load (brown, gc/day) in wastewater samples (panels B, D, F, H, and J). For first test positivity rate, cases, and deaths, dots represent the daily value, and lines the seven-day rolling mean. For N1 gene concentration and daily viral load, dots represent each reading of the samples, and the line was obtained by fitting a LOESS. Five wastewater treatment sites are reported: Levenmouth (A and B), Falkirk (C and D), Dunfermline (E and F), Allanfearn (G and H), and Hamilton (I and J). Vertical lines mark the changes in restrictions: local or minor policy changes (orange dotted lines), the introduction of the regional tier system (dashed red line) and the post-Christmas (26/12/20) national lockdown (black thick line).

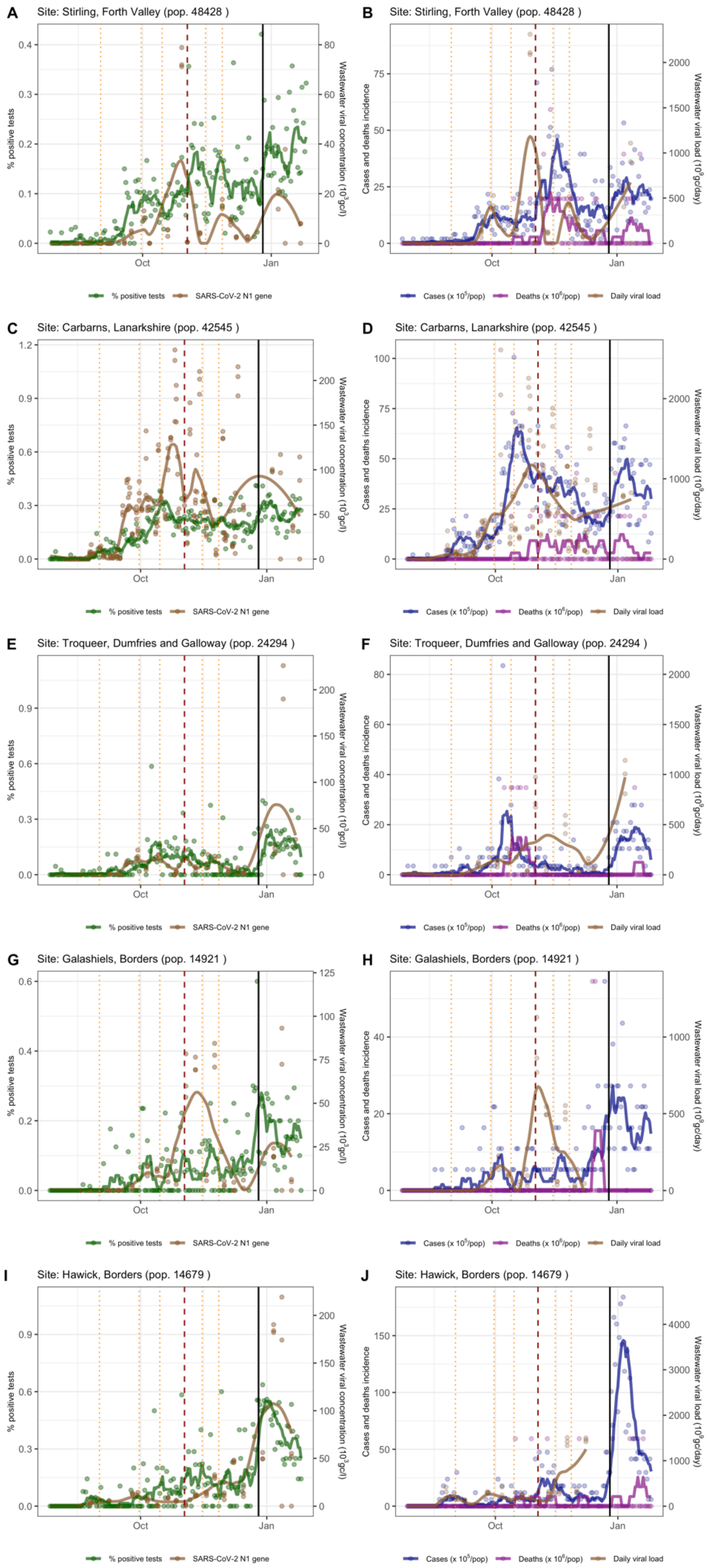


**Figure S2.3**. Trends of the first test positivity rate (green) and SARS-CoV-2 N1 gene concentration (brown, gc/l) in wastewater samples (panels A, C, E, G, and I); trends of incidence per 100’000 people (blue), deaths per 1’000’000 people (purple), and N1 gene daily viral load (brown, gc/day) in wastewater samples (panels B, D, F, H, and J). For first test positivity rate, cases, and deaths, dots represent the daily value, and lines the seven-day rolling mean. For N1 gene concentration and daily viral load, dots represent each reading of the samples, and the line was obtained by fitting a LOESS. Five wastewater treatment sites are reported: Stirling (A and B), Cabarns (C and D), Troqueer (E and F), Galashiels (G and H), and Hawick (I and J). Vertical lines mark the changes in restrictions: local or minor policy changes (orange dotted lines), the introduction of the regional tier system (dashed red line) and the post-Christmas (26/12/20) national lockdown (black thick line).

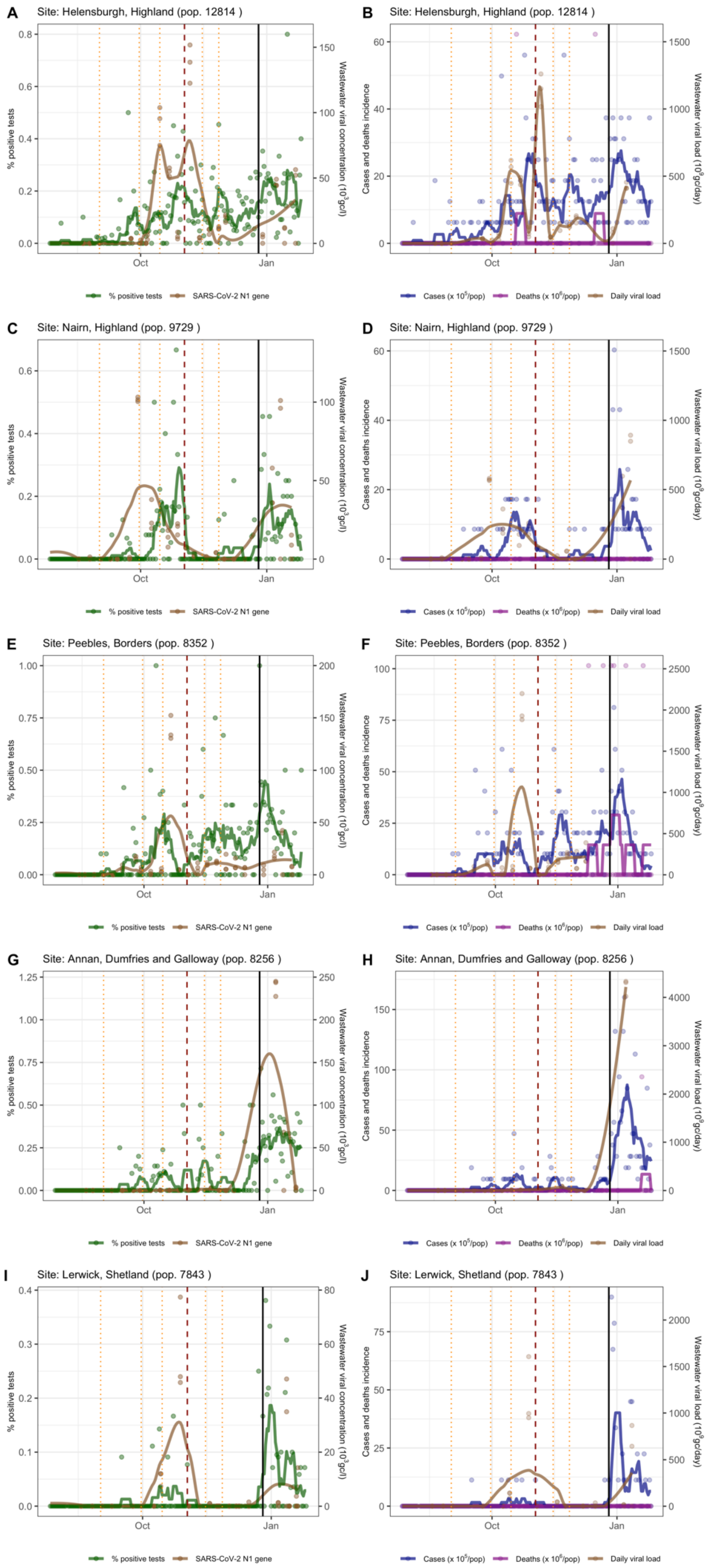


**Figure S2.4**. Trends of the first test positivity rate (green) and SARS-CoV-2 N1 gene concentration (brown, gc/l) in wastewater samples (panels A, C, E, G, and I); trends of incidence per 100,000 people (blue), deaths per 1,000,000 people (purple), and N1 gene daily viral load (brown, gc/day) in wastewater samples (panels B, D, F, H, and J). For first test positivity rate, cases, and deaths, dots represent the daily value, and lines the seven-day rolling mean. For N1 gene concentration and daily viral load, dots represent each reading of the samples, and the line was obtained by fitting a LOESS. Five wastewater treatment sites are reported: Helensburg (A and B), Nairn (C and D), Peebles (E and F), Annan (G and H), and Lerwick (I and J). Vertical lines mark the changes in restrictions: local or minor policy changes (orange dotted lines), the introduction of the regional tier system (dashed red line) and the post-Christmas (26/12/20) national lockdown (black thick line).

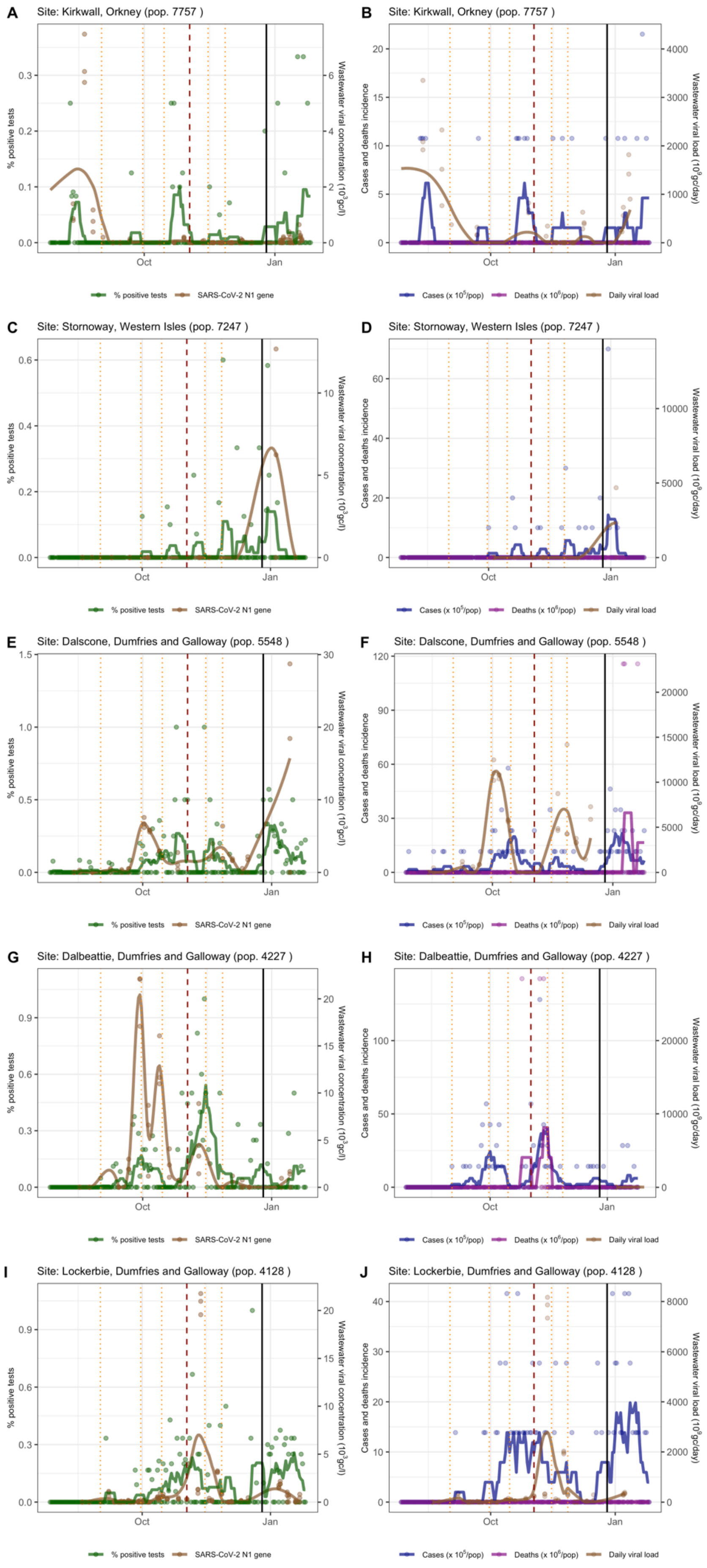


**Figure S2.5**. Trends of the first test positivity rate (green) and SARS-CoV-2 N1 gene concentration (brown, gc/l) in wastewater samples (panels A, C, E, G, and I); trends of incidence per 100,000 people (blue), deaths per 1,000,000 people (purple), and N1 gene daily viral load (brown, gc/day) in wastewater samples (panels B, D, F, H, and J). For first test positivity rate, cases, and deaths, dots represent the daily value, and lines the seven-day rolling mean. For N1 gene concentration and daily viral load, dots represent each reading of the samples, and the line was obtained by fitting a LOESS. Five wastewater treatment sites are reported: Kirkwall (A and B), Stornoway (C and D), Dalscone (E and F), Dalbeattie (G and H), and Lockerbie (I and J). Vertical lines mark the changes in restrictions: local or minor policy changes (orange dotted lines), the introduction of the regional tier system (dashed red line) and the post-Christmas (26/12/20) national lockdown (black thick line).

To estimate the number of SARS-CoV-2 “active shedders” (i.e. the number of infected individuals contributing to the wastewater viral load) for each wastewater sample, we summed the number of positive tests over a specified time window ending on the wastewater sampling day. In the main analysis we report the results corresponding to a seven-day time window, here we test the sensitivity of changing this time window. First, we ran a series of correlation tests (Spearman ρ test), using both the N1 gene concentration expressed in gc/l (viral concentration) and in gc/day (daily viral load, calculated by multiplying the former by the daily flow). These were tested against the number of detected cases and the first test positivity rate in the WWTP catchment area. Results of this correlation are reported in Fig. S2.6 (panel A), which shows how the daily WWTP daily viral load highly correlates with the number of detected cases, while the RNA concentration is better correlated to the first test positivity rate in the catchment area. Our results show how the strength of the correlation increases as the time window increases up to five or six days and then stabilises. When breaking down this correlation by catchment area (Fig. S2.7), we observed that high correlations were maintained across most of the catchments, in particular the larger ones (i.e. WWTPs serving more people). Furthermore, we ran the mixed regression model (see *Data analysis* section in the main text) calculating the number of detected cases for time windows from zero to 28 days. Fig. S2.8 shows the conditional $R^2$ of the model ranging from 0.71 to 0.89, with an average of 0.76. The combination of the correlations and mixed model results suggests that faecal shedding could be mostly concentrated in the few days after symptoms onset, while the contribution to the wastewater of individuals infected for longer might be low or negligible.

Another important consideration is whether the relationship between detected cases and wastewater RNA concentration differs in the phase of the epidemic that precedes the epidemic peak (positive case trend) vs. the phase after the peak (negative case trend). To investigate this further, we divided the observations based on whether samples were taken during the positive or negative trend in cases in the corresponding catchment. This was done by fitting a time series with a weekly “seasonality” (in order to account for the reduced number of tests performed during weekends) and then by extracting the trend. As shown in Fig. S2.6.B, the correlation between daily viral load and detected cases is still very high, independent of the trend in cases. The other correlations were weaker during the negative phase, when compared to the positive phase, although they never dropped below 0.65. Similar results came from two mixed regression models, which we ran by dividing the samples in the same manner (positive vs. negative trend in cases). Here, the conditional $R^2$ was 0.80 during the upward phase, and 0.76 during the downward one.

Given that the WWTP daily viral load is highly correlated with the number of cases in its catchment, it is essential to obtain an estimate of the daily WWTP influent flow in order to calculate the daily viral load from the viral RNA concentration. Since this measure is not readily available for all WWTPs, The Scottish Environment Protection Agency provided an estimate of this value using a simple regression model. As explanatory variables, this model needs the sample ammonium concentration and the catchment population, which can be easily captured for all WWTPs. To check the effect of using an estimated WWTP daily flow instead of the observed one, we re-ran the main mixed model using the estimated WWTP daily flow, instead of the observed flow, to calculate the daily viral load. The impact on the model was minimal, with a conditional $R^2$ of 0.76 i.e. only 0.02 lower than the model parameterised using observed flow.

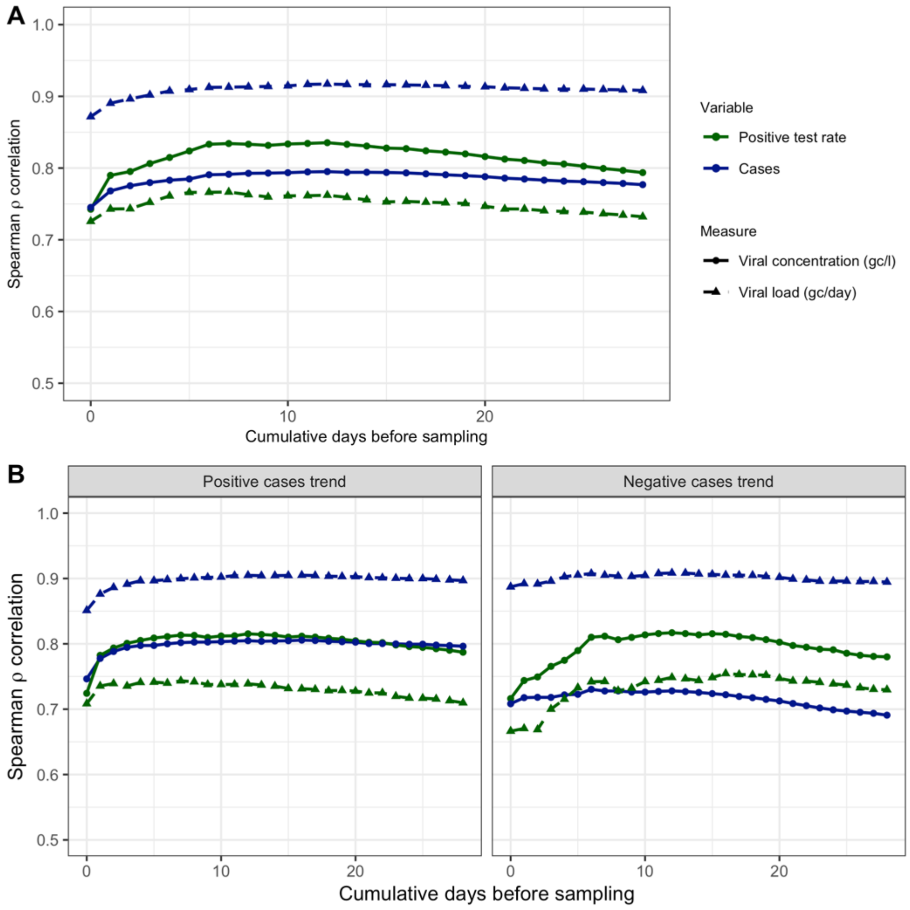


**Figure S2.6**. Spearman correlation between SARS-CoV-2 detected cases and the wastewater viral concentration and load. Detected cases in the WWTP catchment area were summed for a variable number of days prior to wastewater sampling (x axis: 0 to 28 days). Continuous lines and dots represent the values for viral concentration (gene copies/l). Dashed lines and triangles represent the values for the daily viral load (gene copies per day). Wastewater SARS-CoV-2 measurements are correlated against either the number of detected cases (blue) or first test positivity rate (green). A. All observations. B. Observations divided according to the case trend (positive or negative) in the corresponding catchment. All *p* were ~0.

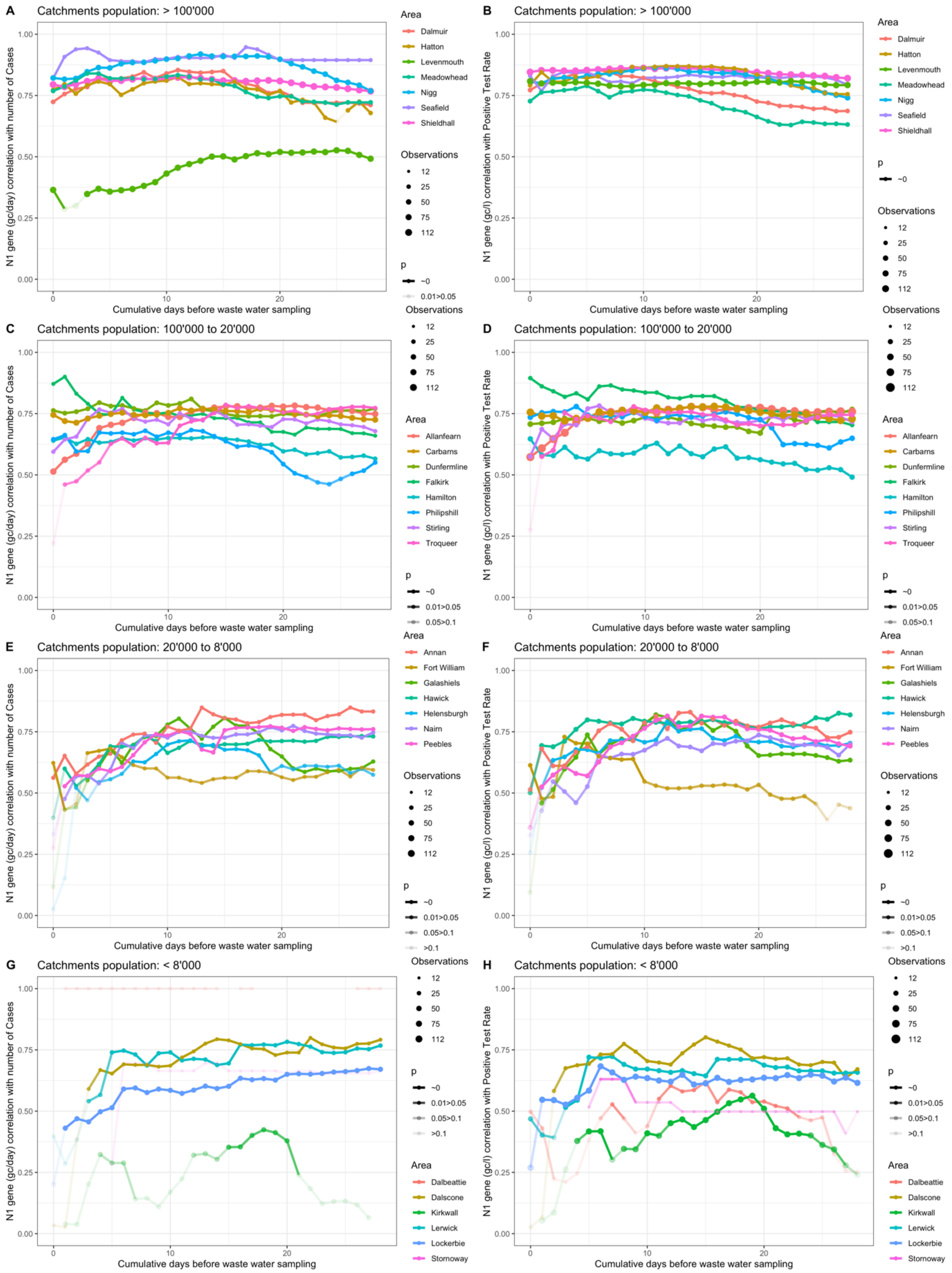


**Figure S2.7**. Spearman correlation between SARS-CoV-2 cases against wastewater daily viral load (panels A, C, E and G); and first test positivity rate against wastewater viral concentration (B, D, F and H). Detected cases in the WWTP catchment area were summed for a variable number of days prior to wastewater sampling (x axis: 0 to 28 days). Wastewater treatment plants are divided by size, and in each plot, colours represent different plants, with dot size corresponding to the number of observations, and line/dot transparency to the *p* value.

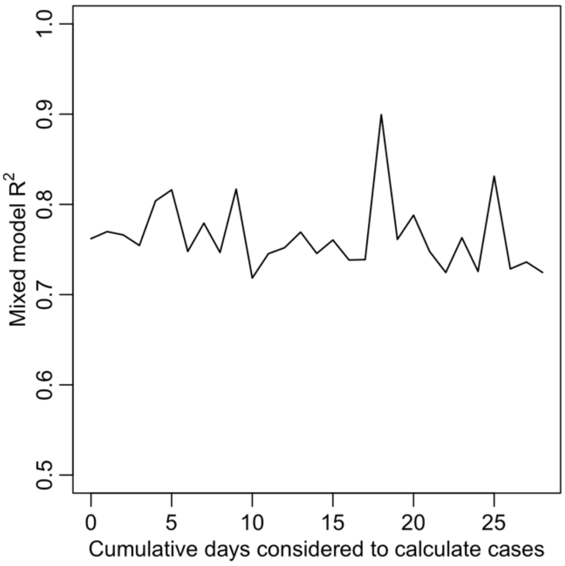


**Figure S2.8**. Mixed regression model performance (reported as the model's conditional $R^2$) by varying the period length over which the number of cases is calculated, from 0 days (i.e. only cases that were recorded on the wastewater sample date are included) to 28 days prior. The $R^2$ varied between 0.71 (10 days) and 0.89 (18 days), and the average was 0.76 (very close to the model reported in the main text, 0.78).

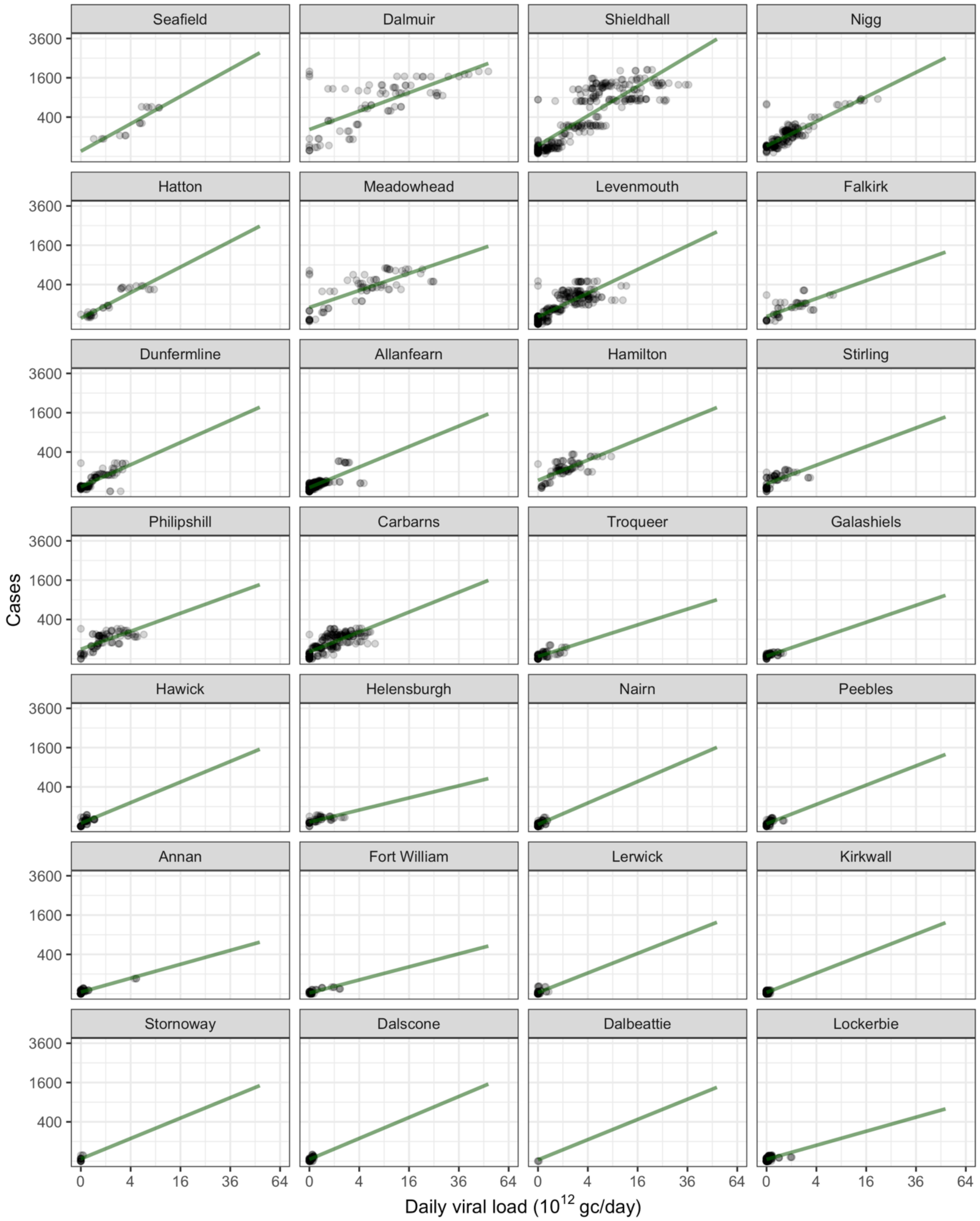


**Figure S2.9**. Linear regression mixed model fit for the 28 wastewater treatment plants, ordered by their catchment population size. In contrast to Fig. 3 (main text), in this figure we fixed the axes scales to visually compare the regression across WWTPs.

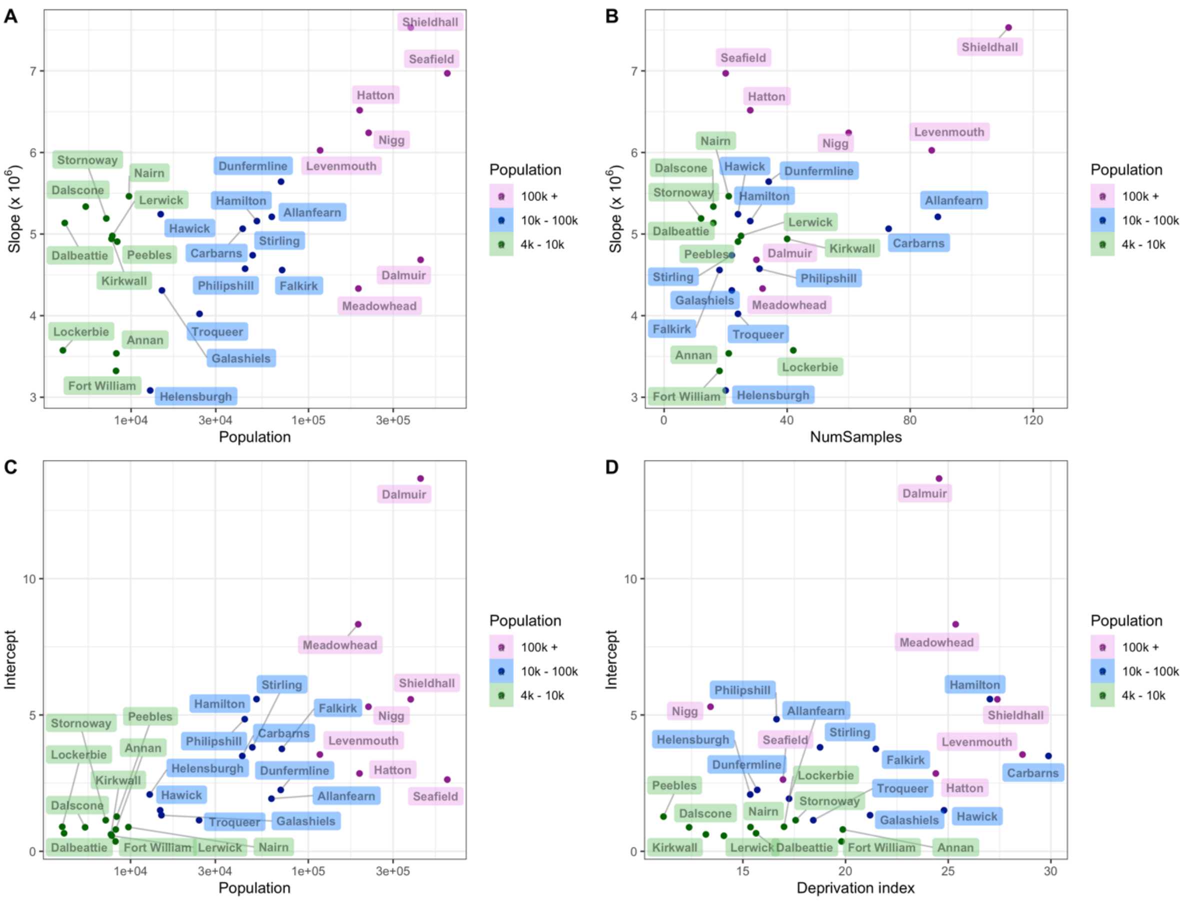


**Figure S2.10**. By using a mixed regression model specifying the sample's WWTP as a random effect, we obtained an estimate of slope (i.e. the effect of the daily viral load) and intercept for each WWTP (n = 28). Here, we plotted these coefficients against the significant variables that explain the differences between WWTPs, previously selected by uni- and multi-variable models. The variables which best explained the coefficients were two: catchment population (panel A) and the number of samples (panel B) for the slope, and population (panel C) and deprivation index (panel D) for the intercept.
